# Unstable periodic orbits are faithful biomarker for the onset of epileptic seizure

**DOI:** 10.1101/2021.09.03.21263098

**Authors:** Mayukha Pal, Sree Bhattacherjee, Prasanta K. Panigrahi

**Author notes:** Corresponding author: Dr. Prasanta K. Panigrahi, Professor, Department of Physical Sciences, Indian Institute of Science Education and Research Kolkata, Mohanpur-741 246, West Bengal, India., Tele.

## Abstract

EEG signals of healthy individuals and epileptic patients, when treated as time series of evolving dynamical systems, are found to display characteristic differences in the behavior of the unstable periodic orbits (UPO), marking the transition from regular periodic variations to self-similar dynamics. The UPO, manifesting as broad resonances in the Fourier power spectra, are quite prominent in their presence in the normal signals and are either absent or considerably weakened with a shift towards lower frequency in the epileptic condition. The weighted average and visibility power computed for the UPO region are found to distinguish epileptic seizure from healthy individuals’ EEG. Remarkably, the unstable periodic motion for healthy ones is well described by damped harmonic motion, the orbits displaying smooth dynamics. In contrast, the epileptic cases show bi-stability and piecewise linear motion for the larger orbits, exhibiting large sudden jumps in the ‘velocity’ (referred to the rate of change of the EEG potentials), characteristically different from the healthy cases, highlighting the efficacy of the UPO as biomarkers. For both the regions, 8-14Hz UPO and 40-45Hz resonance, we used data driven analysis to derive the system dynamics in terms of sinusoidal functions, which reveal the presence of higher harmonics, confirming nonlinearity of the underlying system and leading to quantification of the discernible differences between the healthy and epileptic patients. The gamma wave region in the 40-45Hz range, connecting the conscious and the unconscious states of the brain, reveals well-structured coherence phenomena, in addition to the prominent resonance, which potentially can be used as a biomarker for the epileptic seizure. The wavelet scalogram analysis for both UPO and 40-45Hz region also clearly differentiates the healthy condition from epileptic seizure, confirming the above dynamical picture, depicting the higher harmonic generation, and intermixing of different modes in these two regions of interest.

**Significance:** Unstable periodic orbits are demonstrated as faithful biomarkers for detecting seizure, being prominently present in the Fourier power spectra of the EEG signals of the healthy individuals and either being absent or significantly suppressed for the epileptic cases, showing distinctly different behavior for the unstable orbits, in the two cases. A phase space study, with EEG potential and its rate of change as coordinate and corresponding velocity, clearly delineates the dynamics in healthy and diseased individuals, demonstrating the absence or weakening of UPO, that can be a reliable bio-signature for the epileptic seizure. The phase-space analysis in the gamma region also shows specific signatures in the form of coherent oscillations and higher harmonic generation, further confirmed through wavelet analysis.

## 1. Introduction

Complex dynamical systems in nature are often faithfully modelled by non-linear equations, characterized by stable and unstable periodic orbits, attractors, chaotic behavior, as well as bi-stability, owing their origin to nonlinearity. Human brain is a complex system consisting of billions of nerve cells (or neurons), which help transmit signals from the brain to the rest of the body [1-3]. A seizure is a paroxysmal alteration of neurologic function, arising from the excessive, hyper-synchronous discharge of neurons in the brain. Epilepsy is a serious condition of recurrent, unprovoked seizures. It has numerous causes, each reflecting certain underlying brain dysfunction [4]. Seizures can occur spontaneously as well as in a recurrent pattern, due to distorted neuronal interactions. Epilepsy, being a neurological dysfunction, significantly affects the entire nervous system due to this massive synchronous discharge of the brain cells [5]. Epileptic seizure causes loss of consciousness, sudden jerk movements, muscle spasms, fear, and anxiety, sometimes leading to the loss of life. It is a serious neuronal disorder, with about 1-2% of the global population affected by epilepsy, the most common neurological dysfunction. Epilepsy is not a single disease, but a collection of symptoms that can range from a brief lapse of awareness to prolonged convulsions, all caused by misfiring neurons in the brain. It can be brought on by illness or head trauma, although very often there is no clear precipitating event [6-7].

Electroencephalogram (EEG) recording is a key medical tool for studying the behavior of the seizure. It measures the electric potential of the brain and is capable of providing dynamical neural information, brain disorders, and cognitive processes, related to the brain state [8]. EEG is a non-invasive process and therefore allows the record to be taken for a long period of time for a precise observation of the dynamic behavior of epileptic seizure [9-10]. Brain cells communicate via the electrical impulses through a large number of neurons; its spatiotemporal behavior can be observed by placing about 20 small flat metal discs or electrodes attached to the scalp through wires. This test helps to diagnose medical conditions like epilepsy, head injuries, seizure, brain tumors, and other brain related problems. Sudden seizures due to epilepsy can lead to fatal falls for unattended patients, underscoring the importance of predicting seizure occurrence [11-14]. Difficulty in prediction arises from the lack of reliable biomarkers for detecting the onset of the epileptic seizure [15-17].

The EEG signals have relatively low amplitude levels, from which brain rhythms in the form of waves are well discernible. Commonly observed rhythms include alpha, beta, gamma and theta waves. Alpha waves fall within the range of 4-8 Hz and thus are considered as slow rhythms. These are normal in children up to 13 years and are found in sleeping or in the rest state of adults. Beta waves spike during emotional responses to frustrating events, lying in the range of 13-30 Hz and can be detected on both sides of the head. It is most evident in the frontal domain and gets reduced in case of cortical damage. It can occur during rapid eye movement (REM) sleep. Gamma waves lie within the range of 30-100 Hz and are considered as fast rhythms. These are present during mixed sensory processing such as combination of hearing and seeing. This connects the conscious state to the unconscious state of the brain and is often referred to as genius waves. A reduction in gamma wave activity might be associated with cognitive decline, especially when compared to theta wave activity levels [18-20].

It has been observed that for detecting the epileptic seizure from the EEG data of normal and epileptic subjects, various schemes based on variational mode decomposition [21], wavelet and time frequency analysis [22-25], random matrix theory [26], fractal based analysis [27-29] and neural network and deep learning [30-33] analysis can be potentially used. Here, we report unstable periodic orbits (UPO) as a seizure biomarker from nonlinear dynamics-based analysis. We characterize the complex behavior of this dynamical system, taking recourse to the classical phase-space approach, routinely used in the analysis of electrical circuits, represented by damped driven oscillators. For this purpose, the potential is identified as displacement coordinate with its variation as velocity, whose phase space behavior brings out the characteristic differences. The much-used Fourier spectrum reveals the periodic waves as sharp peaks and self-similar behavior through its scale free power law behavior. These two regimes in the dynamics are often separated by unstable periodic orbits, which appear as broad resonances in the Fourier domain [34]. Remarkably, it is found that the healthy individuals’ EEG signals display well marked UPO, whereas these are absent or significantly reduced in case of the epileptic patients during seizure, signifying its use as a biomarker for the onset of the epileptic seizure. UPO can be regarded as a local signature in the frequency domain for the onset of chaos. The chaotic systems consist of an infinite set of UPO, the trajectory of the system comes occasionally close to a relatively low periodic orbit, resulting in the system to behave visibly in an almost periodic manner for a short time [35-37]. UPO are the “frame” of any dynamical system, which can discern the behavior of a given system. In a chaotic system, they can also take the form of an attractor, appearing like a Lissajous figure. UPO are very sensitive to small perturbations in the environment, as it is located between the boundary of regular and self-similar behavior, characterized respectively by the low-frequency periodic dynamics and higher-frequency chaotic motion. It manifests as a broad resonance, with substantial increase in power in a small range of frequencies in the Fourier domain [38]. Dynamical systems generically possess UPO regions, representing higher frequency periodic motion, that are extremely sensitive to perturbations, making them ideal for the investigation of the characteristically different behavior of the healthy and diseased conditions.

We have observed occurrences of coherent dynamics and also broad UPO, manifesting as resonances, both of which display characteristic differences in normal and epileptic conditions that can act as a biomarker for the onset of epileptic seizure. Focusing on the 40-45Hz gamma region in the Fourier domain, one can identify the differences in the neocortical oscillations, involving most of the cognitive functions in healthy and disease conditions. In the following section 2, the data description is presented along with our analysis methods. Section 3 discusses the results and highlights the UPO biomarkers, with their characteristic differences. Section 4 concludes the study with our inferences and possible future investigations.

## 2. Materials and Methods

This section is devoted to the details of the data that have been used for the subsequent analysis. Methods like the Fourier power spectrum analysis, unstable periodic orbit, phase space plots, and data driven governing system equation derivation are used in this analysis. The measured data is from the extra cranial and intracranial EEG recordings and are used for clinical and research purposes. The data for this work has been taken from the publicly available data of the Department of Clinical Epileptology, University Hospital of Bonn, Germany [39]. The time series data consists of five sets of recordings A, B, C, D, and E, each having 100 channels of 23.6 sec duration, consisting of 4097 recordings. The first and second set measures the extra cranial data of a healthy volunteer in their eyes open and close. The third and fourth set are composed of intracranial recording from hippocampal formation of the opposite hemisphere of the brain of patients and from within the epileptogenic zone during the interictal period, respectively. The data in the last set (E) was recorded during seizure activity (ictal periods) using depth electrodes placed within the epileptogenic zone of the epileptic patients’ brain. The sampling frequency of each data is 173.61Hz. Analysis has been done by subtracting the mean from each single channel initially for each different set and then normalizing it by factoring the standard deviation.

For our analysis, we primarily focused on Set B, the healthy subjects in eye closed condition and Set E, the epilepsy patients during seizure. Analysis for other datasets have been provided in the supplementary material of this manuscript. The Fourier power spectrum is the assignment of power into the frequency components of the signal. It is important in signal processing studies, as it transparently brings out the nature of the different constituents of the time series. The Fourier power spectrum clearly identifies the unstable periodic orbits as broad resonances, as will be shown in our following analysis in the region between 8-14Hz, for the healthy cases. Here the UPO region 8-14 Hz has been taken for the analysis as it possesses the biomarker characteristics because of its sensitive behavior to minute perturbations [34]. The 40-45Hz region has also been taken for consideration as it shows the transition region of higher and lower modulation kind of behavior displaying coherence, as well as a broad resonance.

In our analysis, the phase space trajectories have been generated by plotting the potential values from the recording in the x-axis and the corresponding derivatives in the y-axis. Phase space plotting is a novel way for the study of the EEG data, with the trajectories revealing periodic motion, UPO and sudden changes quite transparently [40]. From the existence of distinct oscillatory patterns of the recorded EEG data, the different brain states can be clearly distinguished [19-20]. Akin to the observations in well studied electrical potential generating circuits, harmonic motion characterized by the following ordinary differential equation, is clearly discernible in phase-space dynamics:

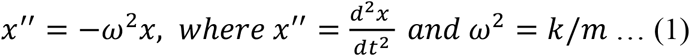

The above is augmented with damping and driving terms, as also non-linear anharmonic terms for appropriate descriptions of diverse physical systems. Our analysis shows some of the trajectories, after following simple harmonic motion, tend to show nonlinear motion like bi-stability and unbounded motion [41]. To ascertain the nature of the nonlinearity affecting the phase space trajectories, we have computed the system behavior in terms of sinusoidal functions and higher harmonics in a data driven approach, which clearly reveals significant presence of higher harmonics, indicative of nonlinearity in the dynamics. One can characterize the different brain waves, alpha, beta, gamma, delta by the occurrence of particular oscillatory patterns from the EEG recordings in different frequency bands. For a time-frequency localization of the bifurcation behavior, as well as intermixing of different modes, we made use of the continuous Morlet wavelet to bring out the transient time varying dynamics in those specific regions of interest: 8-14Hz and 40-45Hz. The oscillatory behavior is analogous to that of harmonic oscillator patterns, originating from Hooke’s law.

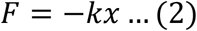

where k is the spring constant, F is the force acting on the oscillator, with x in the form,

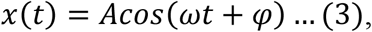

the amplitude A is controlled by the energy of the oscillations, 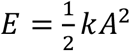.

In physical scenarios, the dynamical behaviors are of two types namely, wave dynamics and particle dynamics. The classical particle dynamics is described by Newtonian mechanics in terms of phase space by considering the coordinate and their instantaneous changes. The wave behavior manifests through coherence effects leading to collapse and revival [19-20]. In the present case, this may arise due to synchronization amongst the neuronal firings. The nonlinear time series from the EEG recordings, when treated as a general dynamical system, can be represented by a general equation of the type.

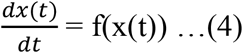

As EEG is recorded during synchronous discharge of electrical activity from the human brain, it is characterized by large fluctuations. The Weierstrass function,

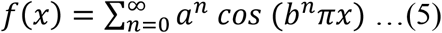

with 0< *a* < 1, b a positive odd integer, is a real valued function known for its continuity everywhere and also is an example of fractal curve, that has been effectively used to characterize biomedical waveforms and complex signals. Fractal geometry has manifested in diverse biological signals and can be potentially useful for the study of the epileptic seizure [28] and can help one to classify and describe the signal at all scales. The nonlinear time series-based investigations have shown positive results in detection and identification of epileptic seizure from EEG recordings. Some of these nonlinear parameters are: Lyapunov exponent [31-32], correlation dimension [31], fractional dimension parameter [28] and approximate entropy (ApEn) [7]. Empirical mode decomposition (EMD) is also a promising method, which helps to develop feature space using ellipse area parameters of two intrinsic mode functions as it is suitable in processing a non-linear time series. The ellipse area parameters of first and second intrinsic mode functions (IMFs) have provided better classification accuracy for classifying ictal and seizure-free EEG signals. EMD method-based decomposition does not require any conditions about the stationarity and linearity of the signal [27]. For identifying a reliable biomarker for the onset of the epileptic seizure, complex behavior of the nonlinear series has been characterized by state of the motion and observed unstable periodic orbits:

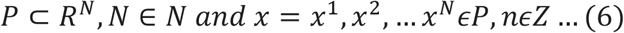

Here P is any number and x is the set of the measurement of a system.

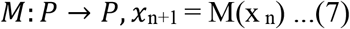

Equation (7) can be, generically, considered as the equation of motion of a dynamical system, where x is the set of measurements of the system, and M is the manifold of the dynamical system.

To understand the dynamics of the phase space structure of the UPO and 40-45Hz region and further corroborate our understanding with the dynamical system’s motion, we make use of a data driven governing system equation derivation approach to derive the system equation. The data used for phase space displacement and velocity plots for the UPO and 40-45 Hz regions are considered for this analysis. Deriving governing equations identifies the physical systems’ linear and nonlinear behavior and hence aids in developing models that can be generalized to predict the future state of the system, the previously unseen behaviors. This is useful in many systems of interest across disciplines where underlying governing equations remain unknown even though a large amount of data is available. It is well known that most physical systems define their dynamics with only a few relevant terms, hence the governing system equations are sparse in a high-dimensional nonlinear function space. So, with use of sparse regression [42-44] and compressed sensing [45-48] combined approach, the dynamical system equation is discovered. The novel approach of combining sparsity methods in dynamical systems brings out system information from data. Here we employed SINDy model [49-51] that considers data x(t) Є ℝ^n^ to discover a best fit dynamical system [52], derived from sparse regression represented by a few possible terms in the form of:

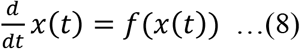

Here the state of the system x evolves with time t with the dynamics being constrained by the function f. To determine the function f from the data, its derivative is either measured or numerically approximated and stacked to form data matrices.

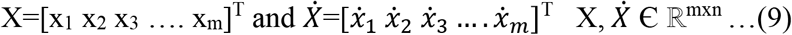

Next, we construct a library consisting of p candidate nonlinear functions from columns of X where ϴ_j_ is a candidate model term and m>>p.

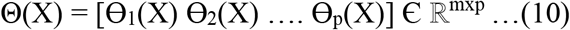

The choice of basis function Θ(X) may consist of constant, polynomial, Fourier and trigonometric terms. Each column of Θ(X) is a candidate function for the right-hand side of Eq. 8. As polynomials are elements of many canonical models hence are most used.

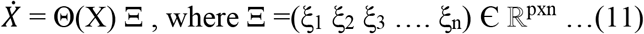

Here matrix Ξ is set of the coefficients which gets the active terms from Θ(X) in the dynamics f. With the use of sparse regression in solving Ξ, each obtained ξ_j_ is sparse with only a few columns selected from Θ(X).

## 3. Results and discussion

The normalized, cumulative sum time series data was considered in Fourier power spectral analysis where the Fourier power is plotted as a function of frequency, in a log-log scale to distinguish the domains of high frequency self-similar behavior from the low frequency periodic ones, with the UPO marking the boundary of these two domains. Comparing the behavior of Set B and Set E, the distinct differences can be ascertained, between the healthy individuals and epileptic patients. Fig 1 represents the Fourier power spectrum (FPS) of the Set B and E, showing the UPO range around 8-14Hz section, centered at 10 Hz. These plots show clear indication that UPO can be considered as a biomarker as here it can be observed that in Set B there is a clear UPO peak whereas it is prominently reduced in case of Set E. Due to the sensitivity of UPO to minor perturbations, it can be considered as a bio-alarm of the onset of the epileptic seizure. In the Supplementary material of the manuscript, we have shown the FPS for Set A. The UPO peaks of Sets A and B are similar, though Set A has recordings with the eyes open, where the role of external stimuli can be important, in contrast to the case of eyes closed, where the healthy volunteer is in a restful state of cognition. Generally, white noise in signal analysis is referred to as a signal with equal intensity at different values of frequencies, resulting in a constant power spectral density. In Fig 1, it can be noticed that the white noise is more prominent in 1(b) than in 1(a), which shows the agitation occurred during the recordings as visible in the FPS analysis.

**Fig 1:**
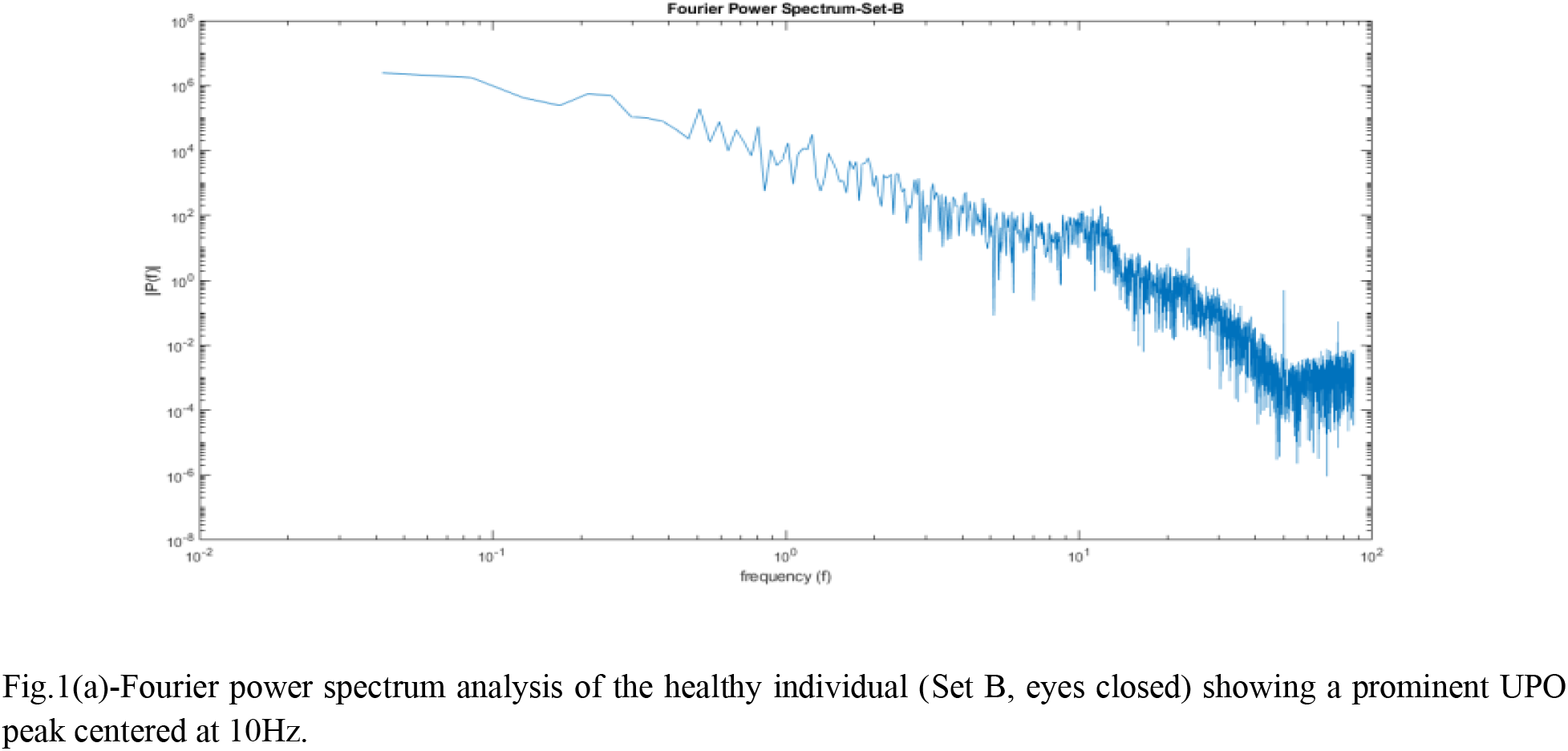
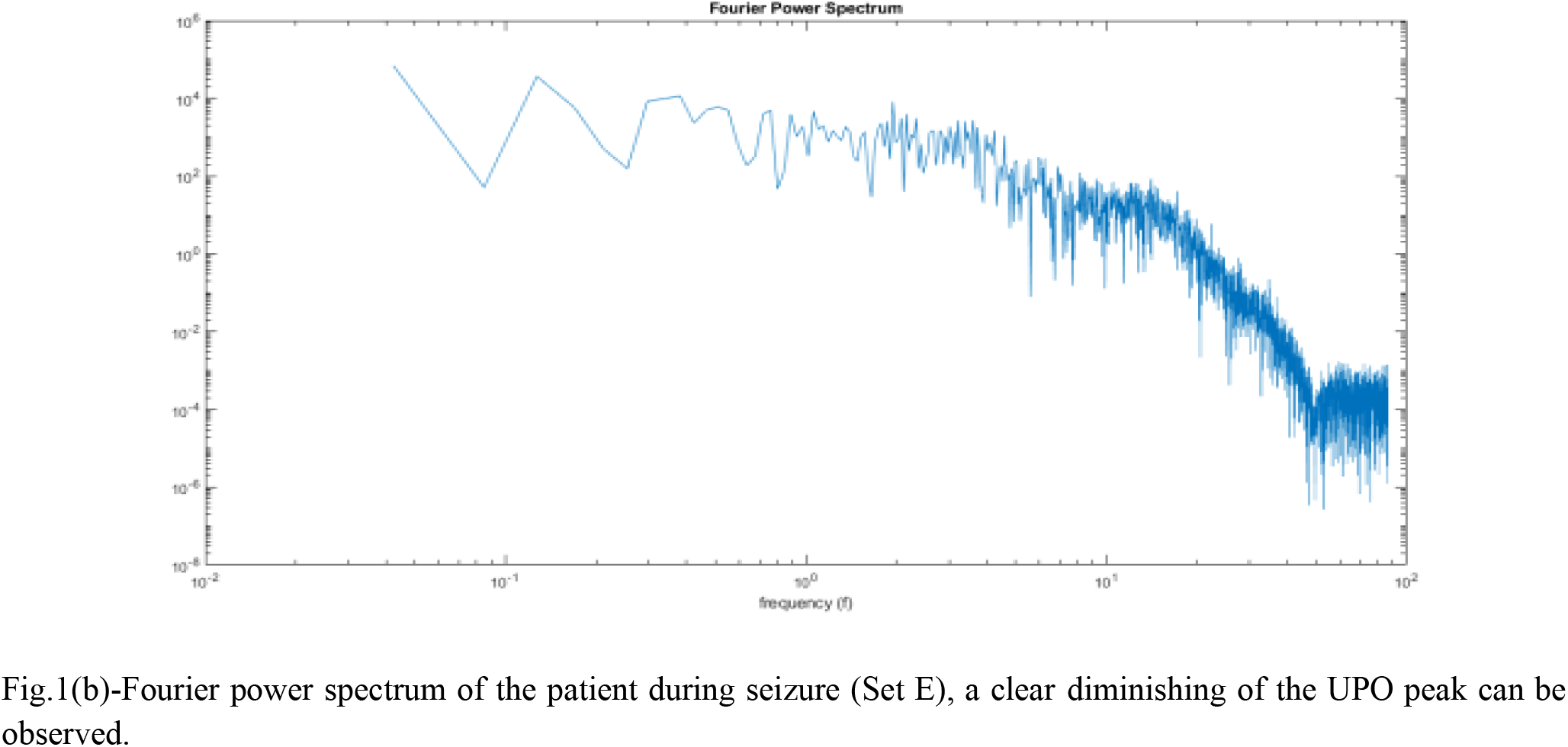
Fourier Power Spectrum for Sets B & E.

We observe dominant UPO regions with gradual increase in power centered around 10-11Hz for healthy subjects, finding diminished power for seizure during the 10-11Hz at ictal activities. Therefore, the quantification of UPO power is a potential biomarker for the seizure activity in epilepsy patients. The UPO region power for set E shows lower power, as compared to healthy subjects, with the power values being random unlike peaks for healthy cases, showing a distinguishing non-harmonic behavior of the EEG waves during seizure activity.

To quantify the UPO power from the FPS for different frequencies from this segment of interest, we have tabulated various data points for both the analyzed datasets. Table 2 depicts the Fourier power for the UPO region of one channel of set B and E to reveal the significant peak power differences. We computed weighted average power for the UPO region to quantify it by calculating the sum product between the frequency and power and then dividing it with the sum of the total frequency. Also, as an additional measure the visibility is computed by subtracting the average of the two neighboring points of the UPO range from the highest power peak value. It is observed from both plots in Fig 2, the quantitative measure of weighted average power and visibility power from the UPO region clearly distinguish healthy individuals from the patients during seizure, reaffirming UPO as potential biomarker. We randomly chose five channels for Set B and E for the representation purpose and verified the features of almost all channels showing a uniform characteristic from UPO. In Set B, all channels the highest peak power is observed at the middle i.e., in and around 11Hz of the UPO region (8-14Hz) with higher power value compared to Set E, where it is at either side of the extreme points of the UPO region with very small or diminishing power. Hence, a one-sided lobe type structure for UPO, with diminishing peaks is observed in case of the epileptic patients during seizure. Also, it is observed that, for both sets E and B, there is oscillatory behavior in the UPO region, both before and after the peak power. Generally, the UPO region from the Fourier power spectrum is detected through measuring the rate of change.

**Table-1:**
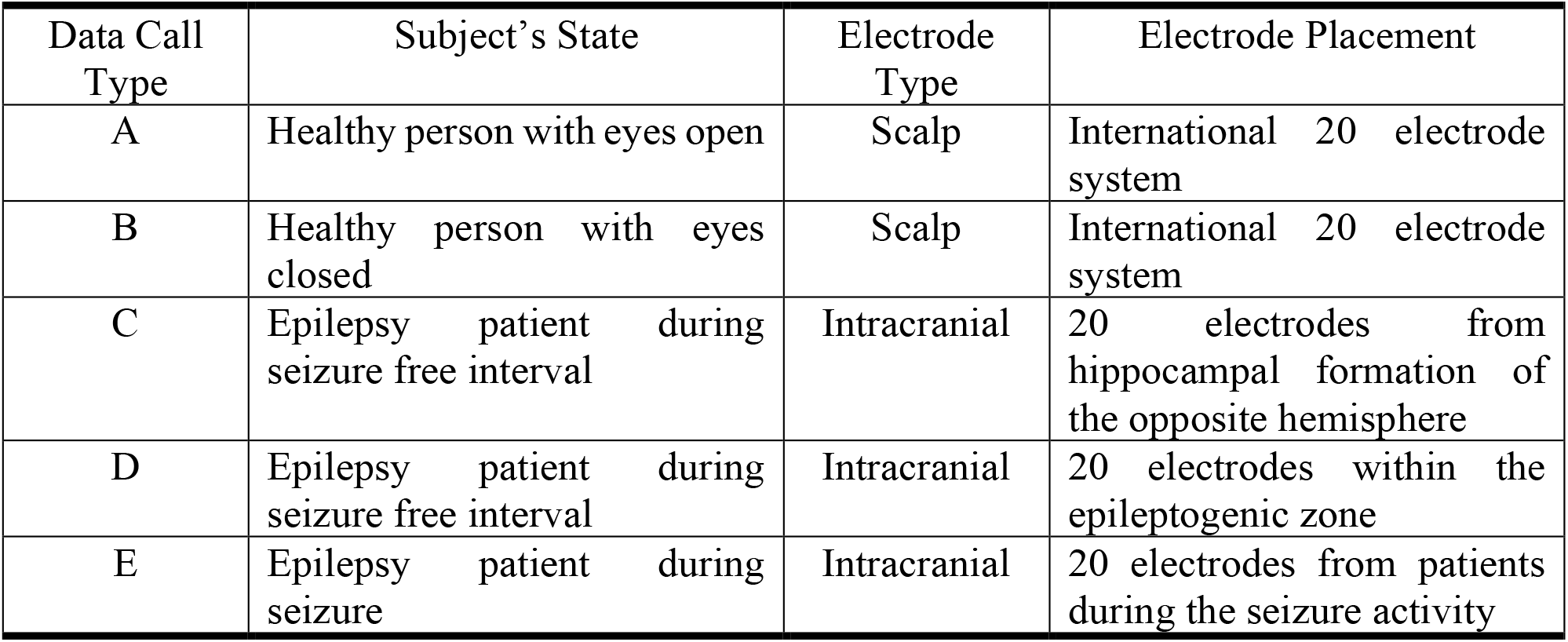
EEG data in brief

**Table 2:** Sample Fourier Power data table for the UPO range of randomly selected channel of Set B (O018) and Set E (S028)

| Set B |  |  | Set E |  |  |
| --- | --- | --- | --- | --- | --- |
| Channel Name | Frequency (f) | Fourier Power (Py) | Channel Name | Frequency (f) | Fourier Power (Py) |
| O018 | 8 | 11.06 | S028 | 8 | 1.876 |
|  | 9 | 14.76 |  | 8.687 | 81.37 |
|  | 10 | 16.88 |  | 10 | 15.89 |
|  | 11.1 | 766 |  | 11 | 51.9 |
|  | 12 | 62.5 |  | 12 | 23.56 |
|  | 13 | 13.18 |  | 13 | 5.422 |
|  | 14 | 4.383 |  | 14 | 8.781 |

**Fig 2:**
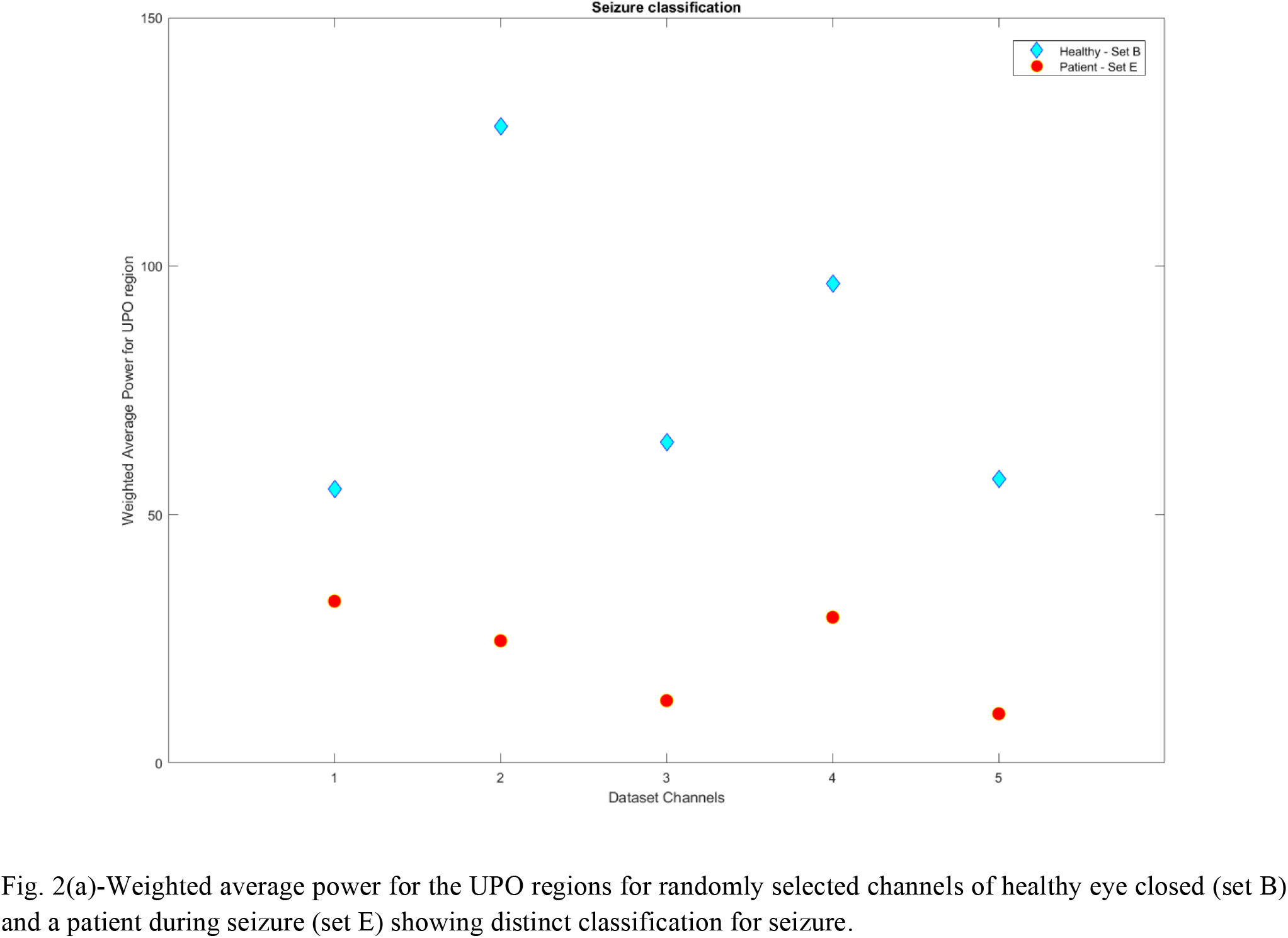
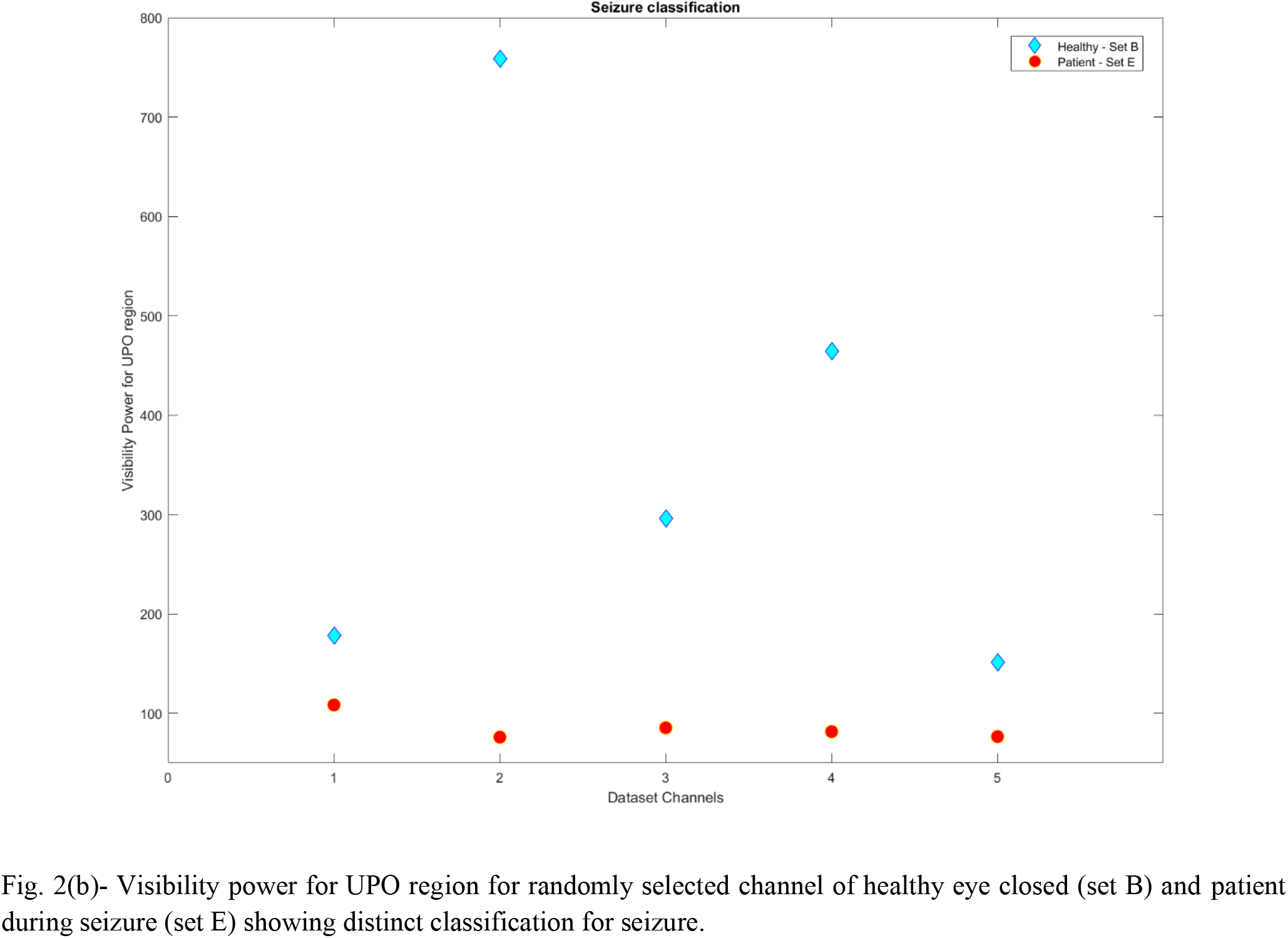
Weighted average and visibility power as quantifying parameter for UPO region to classify seizure.

To understand the nonlinear distribution of power at the UPO region and the 40-45Hz region, we reconstructed the signal from the Fourier Power spectrum only for the UPO or 40-45Hz region. Then the reconstructed signal is used for 2D and 3D scalograms for visualization of UPO as a faithful seizure biomarker and 40-45Hz also has characteristic differences between healthy and seizure patients. Fig 3 and 4 reveal the differences for healthy and seizure patients.

**Fig 3:**
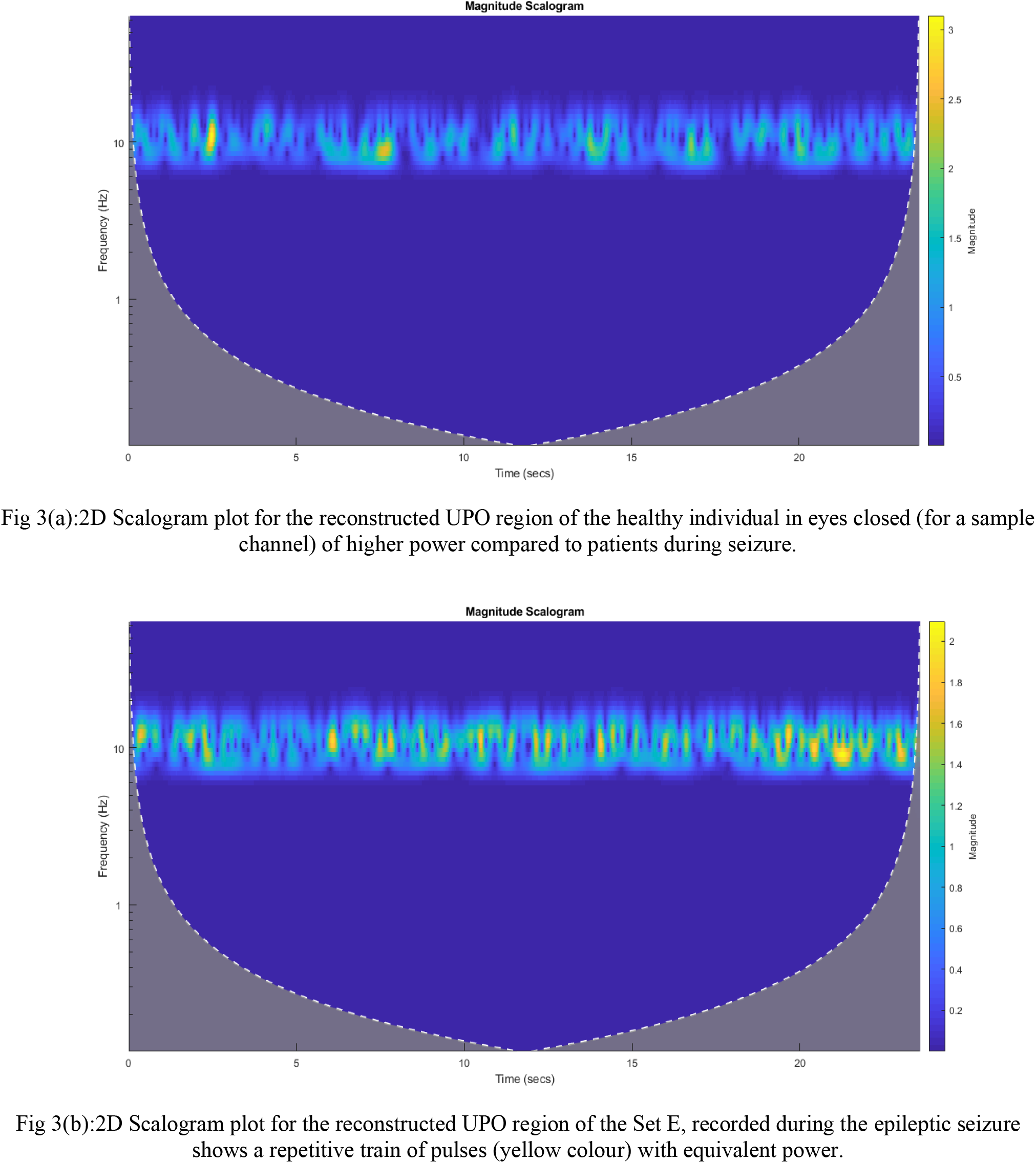
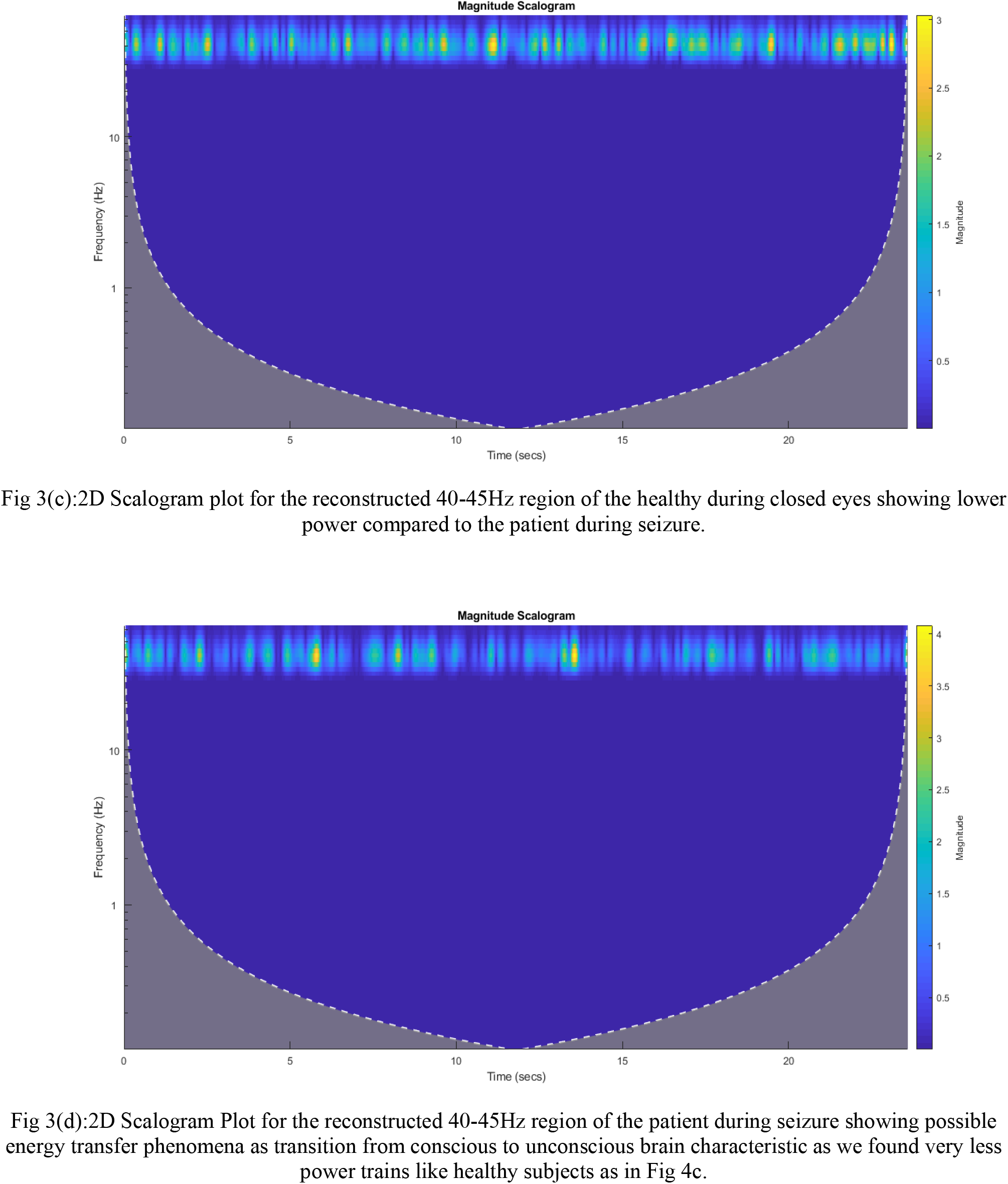
2D scalogram plot of the reconstructed UPO and 40-45Hz region for a randomly selected channel of the Sets B and E.

**Fig 4:**
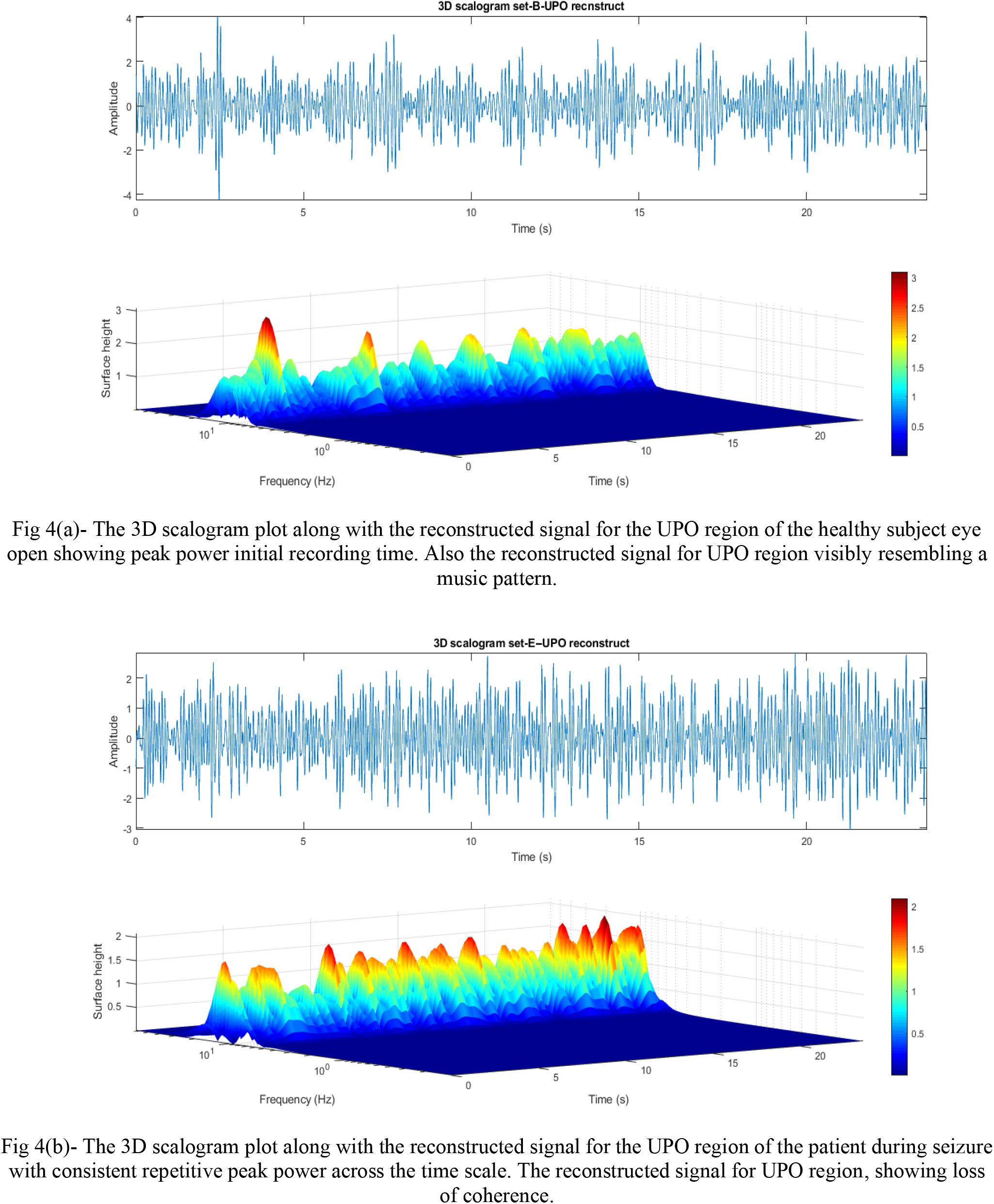
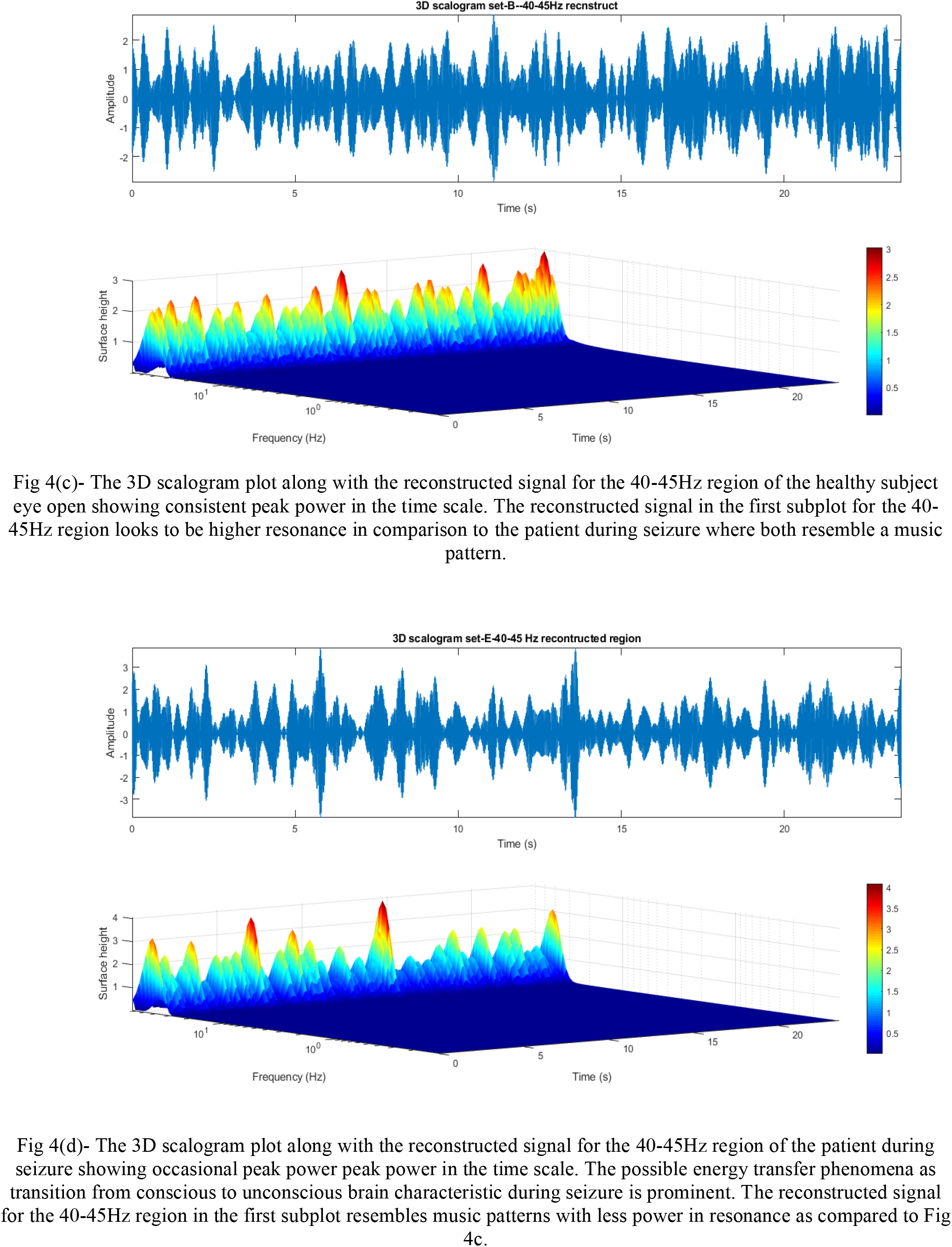
3D plot of the scalogram along with the reconstructed signal for the UPO and 40-45Hz region of the Sets B and E.

### 2D Scalogram Plots

### 3D Scalogram Plots

As we observed bi-stability in phase-space, in our regions of interest, 8-14Hz and 40-45Hz having higher order harmonics in the phase space, along with transient phenomena, we need to study the local properties for this region. To ascertain the time-frequency localization, we use wavelet analysis which reveals temporal behavior for those frequency regions of interest in the data. One clearly observes bimodal patterns in the scalogram plots. The 2D and 3D scalograms along with the reconstructed signal for both UPO and 40-45Hz region as shown in Fig 3 and 4 demonstrate the characteristic difference of scalogram power and reconstructed signal pattern for seizure compared to healthy. To further analyze the dynamics of this UPO region, we have taken the reconstructed time series data from the FPS where except the UPO region we have removed other frequency ranges of Fourier power and reconstructed using inverse Fourier transform. This reconstructed time series obtained from the inverse Fourier transform is used for phase space analysis to understand the dynamical behavior of UPO observed in the analyzed EEG signals. Hence, the phase space diagram emphasizes dynamical activity of the given time-series data. Similarly, we have also studied the phase space structure of the reconstructed signal for the 40-45Hz region.

### UPO Region phase space plots

Further, to characterize these obtained phase structures for the UPO regions of set B and E, we used recently developed data driven discovery of governing physical laws using SINDy approach to derive the system equation for this phase space structure of the UPO. For the analysis, we considered x_0_ as the data from displacement coordinate while x_1_ is the data from velocity coordinate. 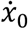 and 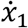 are the derivative of x_0_ and x_1_ respectively used to express the system equations. To correlate with harmonic oscillator motion, we have considered Fourier library with frequency order up to 3 and threshold 0.001 to compute the governing system equations for the set B UPO region phase space structure as shown below:

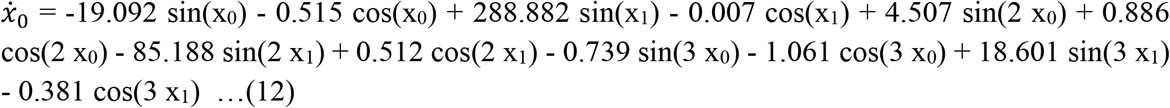

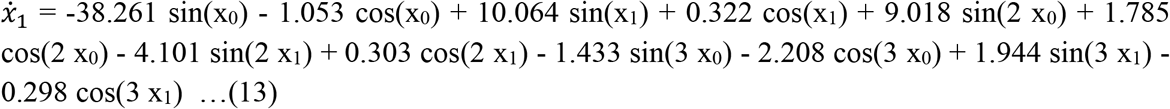

Similarly, for set E UPO, the derived governing system equation in Fourier domain is:

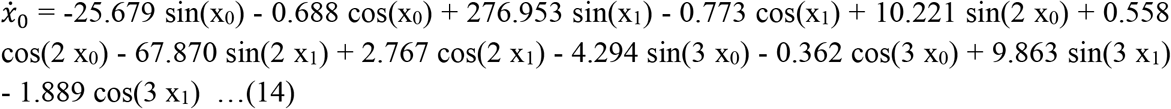

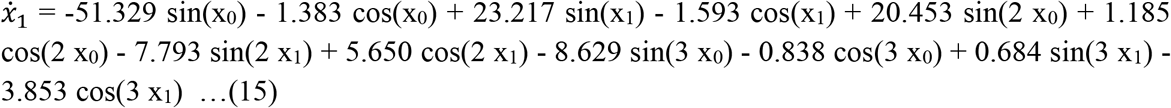

In Fig 5, it is observed that the 8-14Hz UPO region for both the subjects shows simple harmonic motion, thus following Hooke’s law as shown in equation (1), it then gradually changes to non-linear motion. The nonlinear motions generally exhibit quite complicated action over an extended time interval, like chaos. Sometimes under restricted conditions, linear differential equations appear as the approximations to nonlinear equations. The plots are also depicting bimodal behavior; thus, they have two stable shapes, and the multiple trajectories are orbiting around it in periodic motion and also showing unbounded motion like that of an Impact oscillator [53]. On comparing the two sets it has been observed that the bi-stability is stronger for the patient during seizure than that of the healthy subject in stable state. It is well known that the low dimensional chaos is a characteristic of many physiological oscillatory systems including the brain. Time series EEG data in a stable state has been analyzed in a frame of nonlinear dynamics like the attractor dimension. UPO here reveals its efficacy as a faithful biomarker for seizure state with its diminishing power. On careful observation, these plots show similarity to the well-known ‘Lissajous’ figures [52]. In Set E ictal period recordings where data is collected from a depth electrode, it is observed to possess a higher contribution of potential energy, manifesting through a denser plot and the bimodal or the bi-stability gets stronger in comparison to the healthy subject. In the plots the central dense region shows the simple harmonic behavior of the concentric ellipses; they then gradually transition to non-linear unbound type of motion similar to the impact oscillator. An impact oscillator is a periodically forced system that hits an obstruction whenever the displacement reaches a threshold value and a number of continuous impacts originate when the maximum displacement value of regular oscillatory motion equals this value, such events are known as grazing bifurcation [15]. In the phase-space plots it could be observed that the displacement-velocity and velocity-acceleration plots as shown in supplementary material of this manuscript also shows similar kind of behavior as we know that of the harmonic equation.

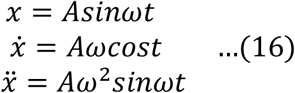

**Fig 5:**
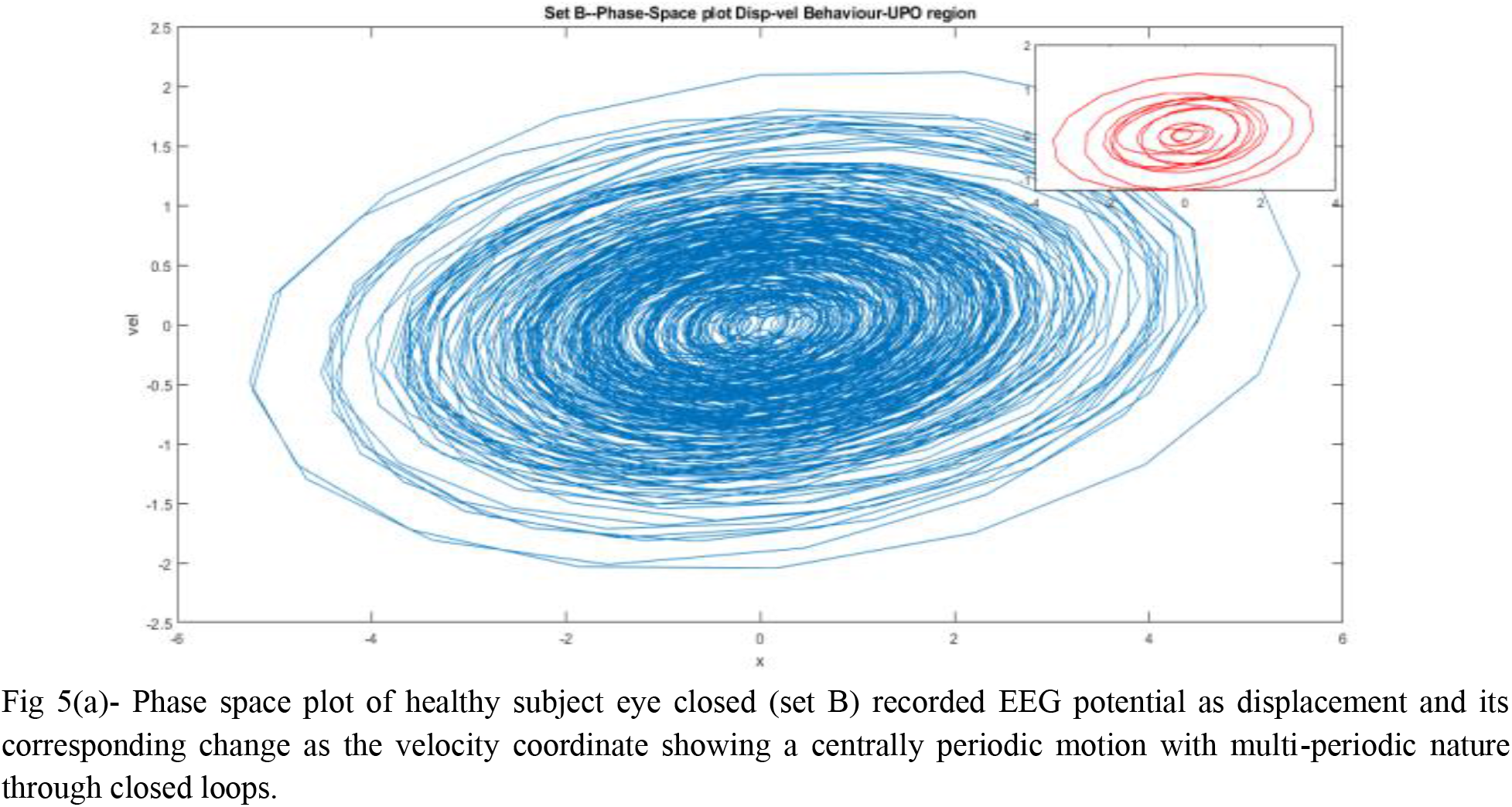
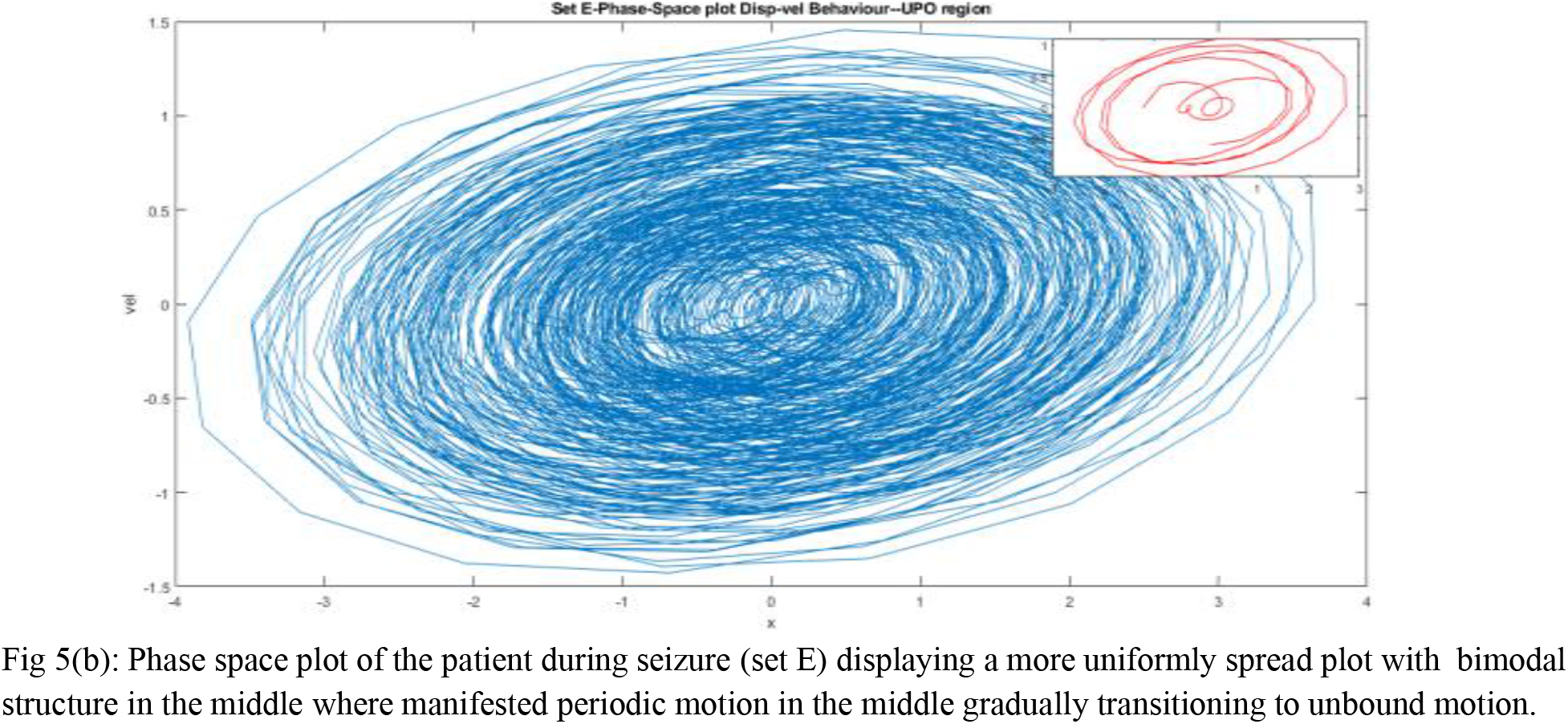
UPO region displacement-velocity phase space plot.

The derived governing system equations in Fourier form from the data for these UPO region phase space structures of both subjects show similar harmonic oscillator system description as in equation 16. Thus, in the displacement-velocity behavior we are getting the relation between sine and cosine terms, hence resembling these plots. The higher order terms in sine and cosine functions in the obtained governing system equation points to the nonlinearity present in both UPO and 40-45Hz, instead of a simple harmonic oscillator. The nonlinearity brings in anharmonicity to the simple damped driven oscillators leading to bi-stability, higher harmonic generation and limit cycle behavior [9, 54]. It is worth emphasizing that we observe the characteristic impact oscillator behavior, when a grazing mechanism leads a non impacting periodic orbit to bifurcate into the impacting one [16]. Like the duffing oscillator the outer orbits tend to return back to the origin which is again matching with the nonlinear nature of the impact oscillator. These faithful reproductions of physical behavior from the EEG UPO region analysis confirm the previous observations and its use as biomarker for medical usages.

### 40-45Hz region phase space plots

Similarly, with use of SINDy model for set B 40-45Hz region reconstructed data and the derived governing system equation in Fourier domain is:

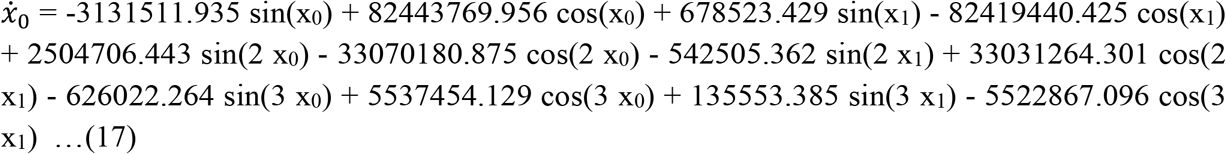

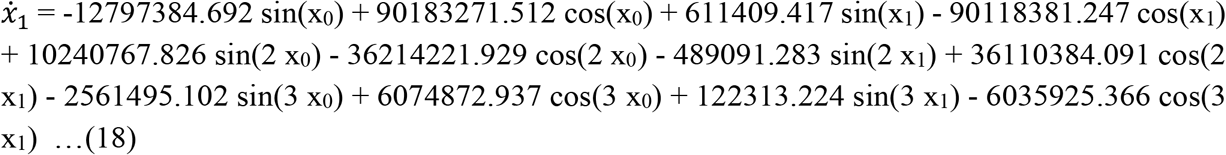

Whereas system equation for set E 40-45Hz region phase space structure is:

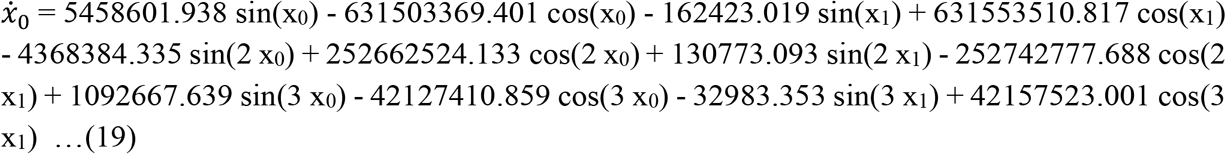

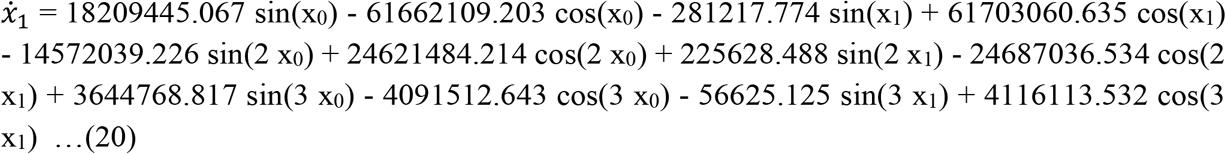

From the above 40-45Hz phase space plots as in Fig 6, it can be concluded the occurrence of the energy transfer phenomenon [19] with conservation of energy states here. In this 40-45 Hz region the coherence phenomenon gets more intense and in case of the epileptic patient plot (epileptogenic zone and during seizure) the potential energy is seen to be more dominant (thus the kinetic energy decrease) than in the case of the healthy subjects. The velocity, acceleration plots for the 40-45Hz region are shown in the supplementary material section of this manuscript where near the central region it separates out the lower value voltages compared to the higher value voltage. The small oscillations are having linear motion, but the large oscillations are tending to acquire an unbound kind of motion like an impact oscillator.

**Fig 6:**
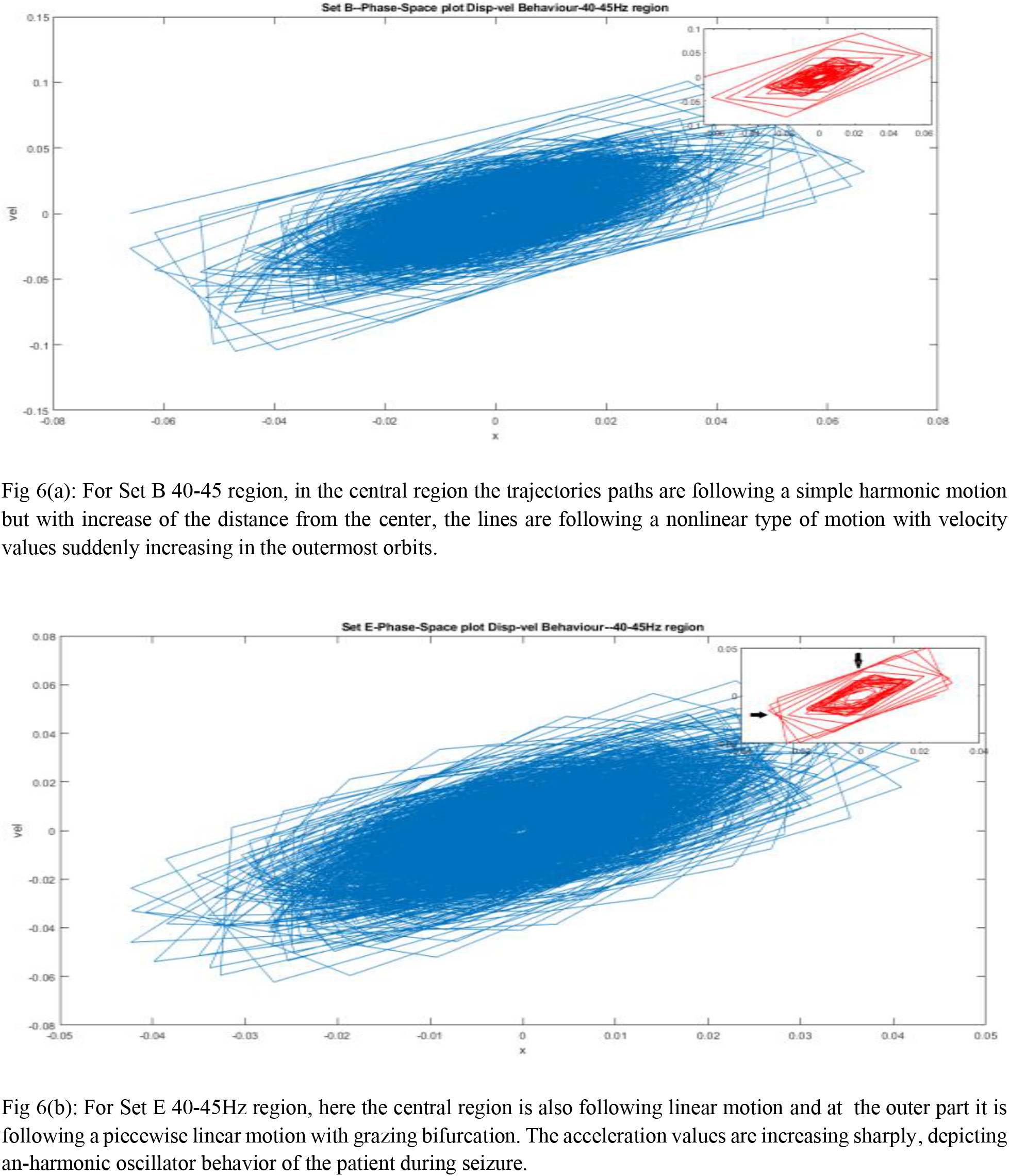
40-45Hz region phase space displacement-velocity plot.

In Fig 6(b), it is seen that the inset figure of 100 data points with the marked black arrows show where acceleration or the velocity values are zero, clearly revealing that the region possesses minimum kinetic region as well as the maximum potential energy. If we observe over the plot the crowd in those points are clearly visible whereas in the inset figure it is only showing the emerging feature which itself has great importance in this analysis, because it shows the local coordinates 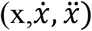 characteristically similar behavior as of the plot which is not possible in case of wave features (with sine and cosine). Thus, we could also consider this energy transition phenomena at 40-45Hz section as a biomarker for the onset of the epileptic seizure. From the plots it is confirmed that a small perturbation in the potential value results into a significant variation in the corresponding velocities, or its rate of change in a piecewise linear motion in the extreme orbits which is indicative of grazing impact oscillator type dynamical characteristics, of non-linearity. It is also seen, if we consider the plots for Set E and Set D (the intracranial epileptogenic zone recordings for the patient shown in supplementary material section) it is observed that the plots for these two sets shows similar behavior in the sense of the path, trajectories but the contribution of potential energy is higher in Set D, which results in a similar structure as each other, which is quite expected because both are from the patient during ictal and inter- ictal period. Other phase space plots for the datasets are given in the supplementary material of the manuscript.

Hence the phase space dynamics help as a bio-alarm for the seizure in epileptic patient. Our derived governing system equation for 40-45Hz from the phase space data complements the analysis. In Table 3, we show the coefficient amplitudes of all sine terms from the governing system equations to give a comparative view of the nonlinearity terms and it’s spread for the UPO and 40-45Hz region. From the equation in table 3, we observe the higher order sine term has lesser amplitude as compared to lower order one relating to the higher order nonlinear phenomena existence. We observe such nonlinear phenomena by the weaker presence of higher order sinusoidal terms for the UPO region, as compared to the 40-45Hz range, which points to the high order nonlinearity at 40-45Hz relating energy transfer phenomena from conscious to unconscious brain state. Also, it is observed that Set B 40-45Hz higher order sine terms mostly have lower amplitude, as compared to Set E which points to the nonlinearity of higher order in patients during seizure compared to healthy subjects. We also observe from the equations that the sine term amplitudes are generally higher than that of the cosine terms for the UPO region, whereas the cosine terms have higher magnitude for the 40-45Hz region compared to sine terms. This corroborates the energy transfer phenomenon occurring in the 40-45Hz region as it is observed to be having higher coherence phenomena where the brain state transitions from the conscious to the unconscious state. In Fig. 7, we show the derived governing system equation for the reconstructed signal used in phase space orbit closely matches with the data. The derived governing equations could be used to predict the future state of the system hence enables here prediction of the future phase space trajectory evolution at UPO and 40-45Hz region that allows easy identification of change in physical/biological phenomenon in these regions.

**Table 3:** Amplitude of the coefficients of all sine terms in the governing system equations of both UPO and 40-45Hz region for healthy and patients. Sine amplitudes of x_1_ are found to be higher than x_0_ and the 40-45Hz amplitudes are higher compared to UPO amplitude.

| Sl.No. | Set Name | Region | Var. | Amplitudes (Coefficients) |  |  |  |  |  |
| --- | --- | --- | --- | --- | --- | --- | --- | --- | --- |
| | | | | Sin($x_0$) | Sin($x_1$) | Sin($2x_0$) | Sin($2x_1$) | Sin($3x_0$) | Sin($3x_1$) |
| 1. | Set-B | UPO | $\dot{x}_0$ | -19.092 | 288.882 | 4.507 | -85.188 | -0.739 | 18.601 |
| | | | $\dot{x}_1$ | -38.261 | 10.064 | 9.018 | -4.010 | -1.433 | 1.944 |
| | | 40-45 Hz | $\dot{x}_0$ | -3131511.935 | 678523.429 | 2504706.443 | -542505.362 | -626022.264 | 135553.385 |
| | | | $\dot{x}_1$ | -12797384.692 | 611409.417 | 1240767.826 | -489091.283 | -2561495.102 | 122313.224 |
| 2. | Set-E | UPO | $\dot{x}_0$ | -25.679 | 276.953 | 10.221 | -67.870 | -4.294 | 9.863 |
| | | | $x_1$ | -51.329 | 23.217 | 20.453 | -7.793 | -8.629 | 0.684 |
| | | 40-45 Hz | $\dot{x}_0$ | 5458601.938 | -162423.019 | -4368384.335 | 130773.093 | 1092667.639 | -32983.353 |
| | | | $x_1$ | 18209445.067 | -281217.774 | 14572039.226 | 225628.488 | 3644768.817 | -56625.125 |

**Fig 7:**
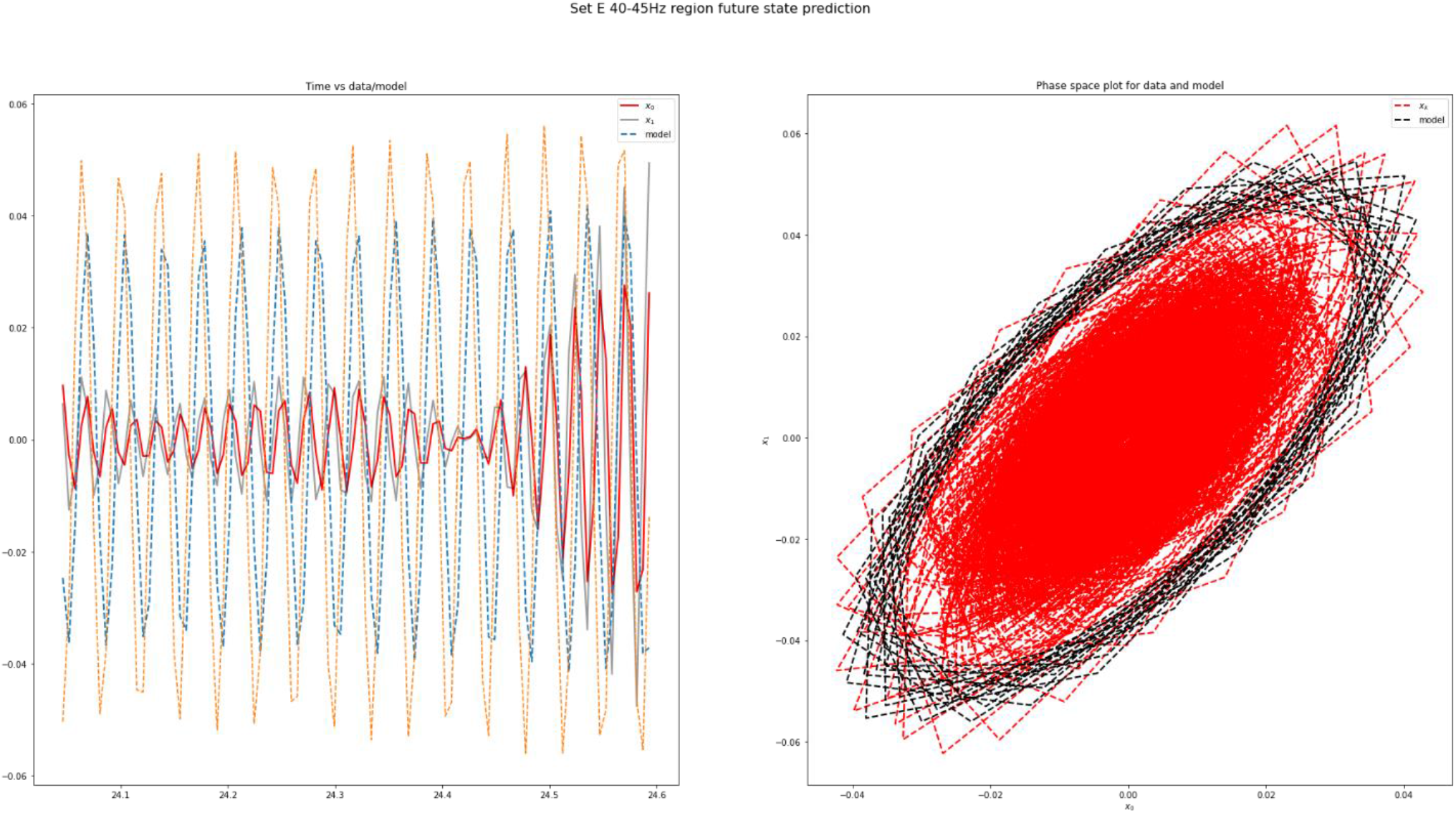
Plot of displacement-velocity phase space data along with the model data for set E(patient during seizure) 40-45Hz region. We observe the derived governing equation/model matches to the phase space displacement-velocity data in the first subplot where x-axis is time, and in the second subplot phase space is drawn for data and the model that enables future phase space trajectory state prediction. Other plots for set B for 40-45Hz and UPO region are shown in the supplementary material of the manuscript.

## 4. Inference

The Fourier power spectrum analysis to identify the UPO region and further its reconstructed signal from inverse Fourier transform to use in the phase-space analysis for different subject EEG recordings of the brain can be used for finding the biosignature on the onset of epileptic seizure. As brain signals are from a nonlinear dynamical system whose behavior is complex hence it is highly effective for the analysis of epilepsy. The bi-stability in the plots show nonlinear behavior of the data. Fourier spectral analysis method allow us to separate the UPO domain precisely in phase space perspective. Together with a data driven approach, the governing system equations were derived to corroborate the understanding of nonlinearity observed from the phase space orbits which also allows future state prediction of phase space trajectory for UPO and 40-45Hz region. The Newtonian mechanics approach in terms of potential and kinetic energy difference and differing grazing orbits often possess piecewise linear motion. For the case of the brain signals we observe impact oscillator type behavior and symmetries in phase space, originating from coalescing of the orbits. The observed coherence and higher order nonlinearity at 40-45Hz region corroborates the understanding of energy transfer phenomena and effectively the brain state transition from conscious to unconscious state with characteristic difference between healthy and seizure. The time-frequency localization using wavelet analysis to understand the transient phenomena at these region of interest 8-14Hz and 40-45Hz shows bimodal patterns with distinct behavior for healthy and seizure. Potentially, 40-45Hz could also be used as biomarker for seizure and we wish to focus our study on future. As epilepsy is a serious disease affecting a considerable amount of the global population hence these investigations may find its significance in biomedicine.

## Supporting information

suppl mat

## Data Availability

The obtained data for our analysis is from free public database published by Department of Clinical Epileptology University Hospital of Bonn Germany [39]. As per the reference [39] the original data collection methods were carried out in accordance with relevant guidelines and regulations where all experimental protocols were approved by the University of Bonn ethics and licensing committee and written consents were obtained from all subjects by the author in ref [39] to make the data available in public for further research.

http://www.epileptologie-bonn.de/cms/upload/workgroup/lehnertz/eegdata.html

## Appendix A. Supplementary material

Supplementary material to this article can be found with this manuscript.

## Acknowledgements

This research work did not receive any specific grant from funding agencies in the public, commercial, or not-for-profit sectors. The author MPal wishes to thank ABB Ability Innovation Center, Hyderabad for their support in research work. The authors alone are responsible for the content and writing of the paper.

