## Supplementary material for "Unstable periodic orbits are faithful biomarker for the onset of epileptic seizure": suppl mat

### Supplementary Materials:

The brain EEG signal on eye open (A) and eye close (B) conditions shows a difference, the former gets perturbed by the external stimuli due to opened eye, whereas the latter is free from it due to eyes in closed condition. The signals from the patient from the epileptogenic zone (D) and during seizure show similarities in their behavior, while the differentiation is characterized as it can indicate the signature prior to start of the seizure. It has been well established that the potentials originate from a source, effectively represented by a localized LCR unit, which in brain  neurons can synchronize with others to generate alpha, beta, gamma waves or individual signals which can be chaotic. In search of biomarker, we found UPO and 40-45Hz has given good results in characterizing epileptic seizure form healthy subjects. We have shared results of various figs that could not be accommodated in the manuscript as additional supporting material to our findings.

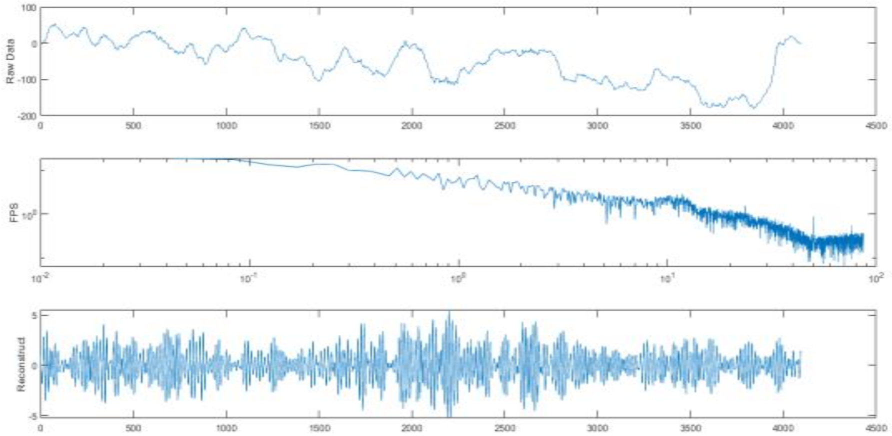


Fig 1(a): Reconstructed UPO signal for the healthy  Subject (eyes close), with the first subplot showing raw data where plot looks periodic, the third subplot is displaying the reconstructed signal for the UPO region with strong oscillatory regions.

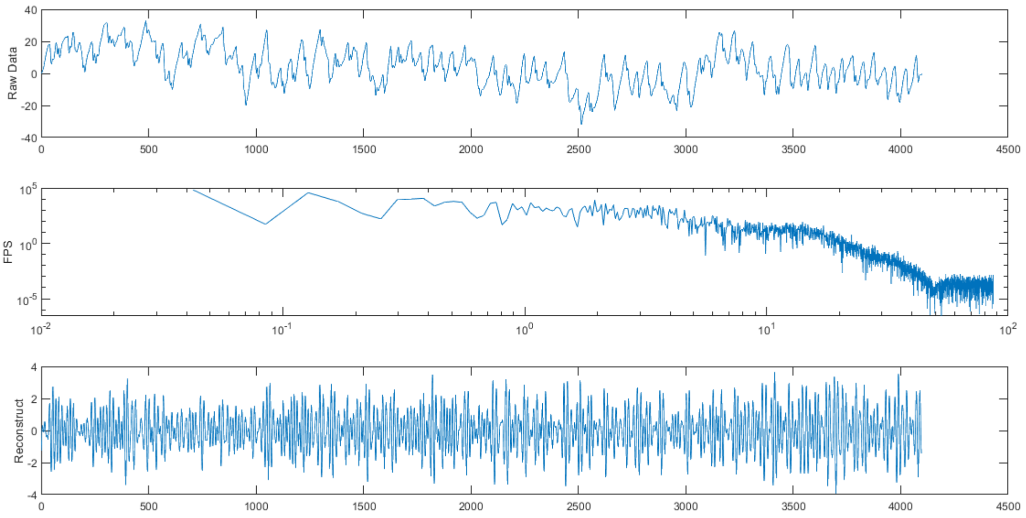


Fig 1(b): Reconstructed UPO signal for  the patient during ictal period the first subplot shows a highly fluctuating plot for the raw data, and reconstructed plot is displaying high amplitudes but with reduced oscillatory region

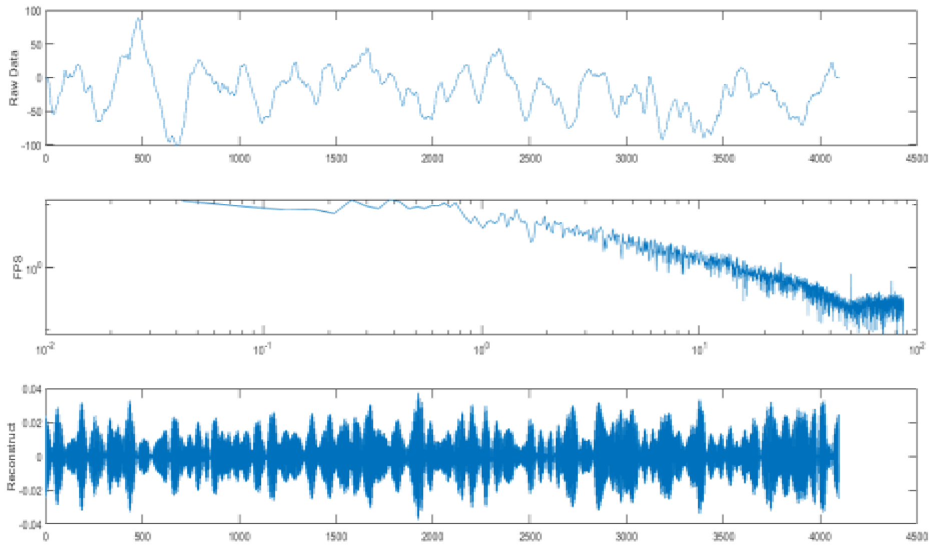


Fig 1(c)- Reconstructed signal of the region 40-45Hz for healthy eyes closed showing stronger coherence than that of the UPO region.

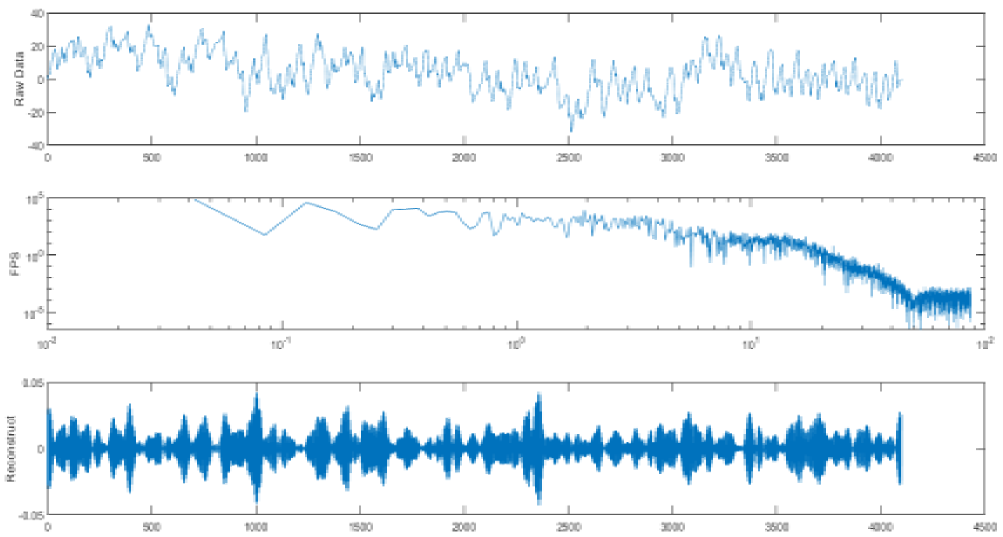


Fig 1(d)- Reconstructed signal of the 40-45Hz region for the patient during seizure showing stronger coherence behavior and faster superposition of the waves during compared to UPO region for same subject.

**Fig 1:** Reconstructed signal from Fourier Power spectra for the UPO and 40-45 Hz region.

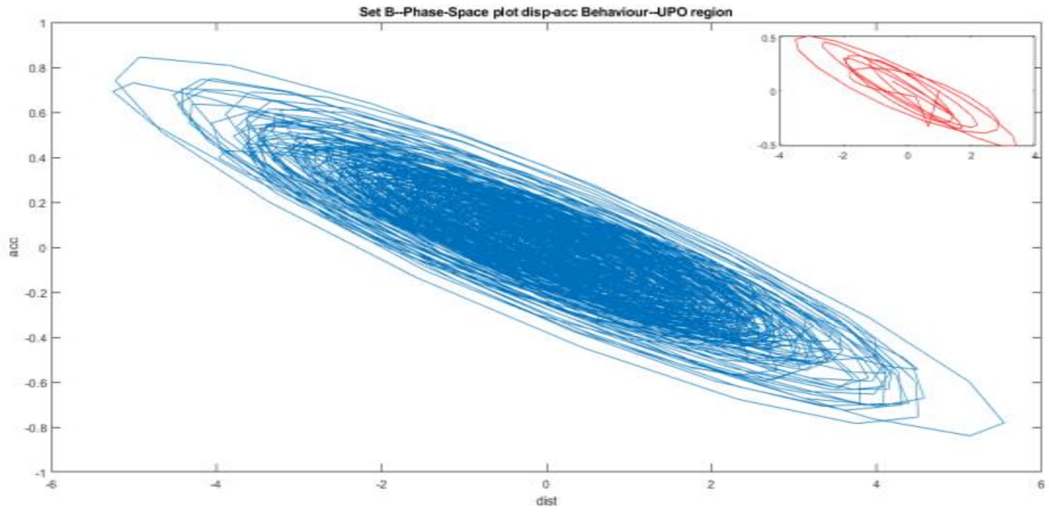


Fig 2(a)-Phase-space plot of displacement and acceleration for healthy individual (eyes closed) UPO region showing periodic behavior then gradually obeying a non-linear to the unbounded motion.

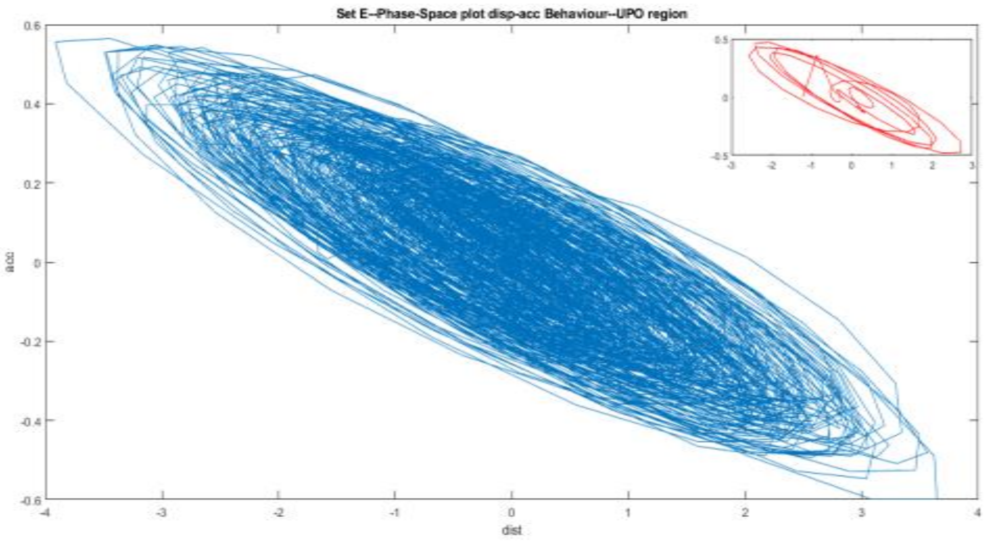


Fig 2(b)- Phase space plot of displacement and acceleration for the patient during seizure at UPO region showing a denser loop all over than that of healthy subject before following the unbounded motion.

**Fig 2:** Displacement-Acceleration phase space behavior for UPO region for both healthy and seizure.

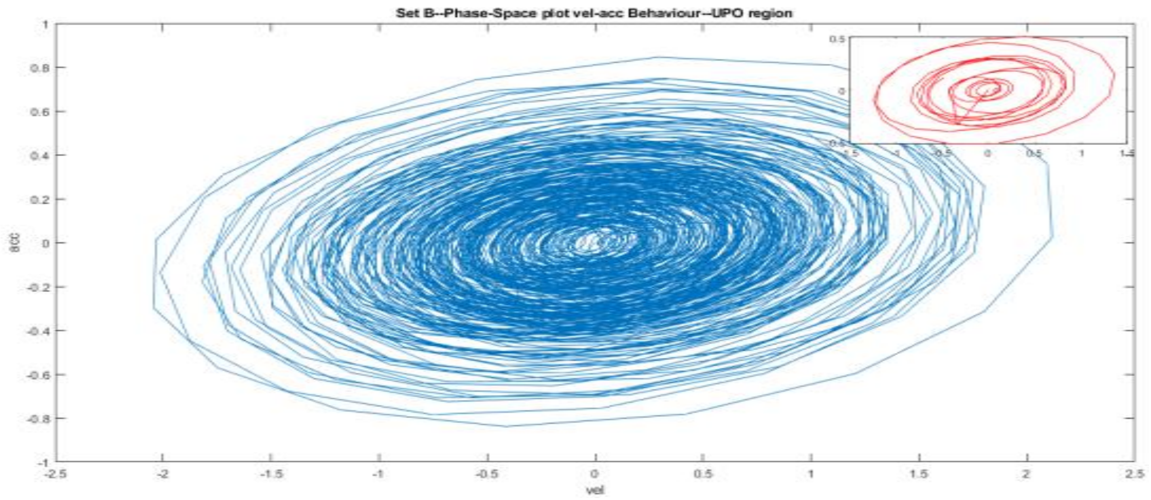


Fig 3(a)- Phase space plot of velocity – acceleration for healthy person with eyes closed for the UPO region showing bimodal structure at the centre with single particle like oscillation having closed orbits of varying size.

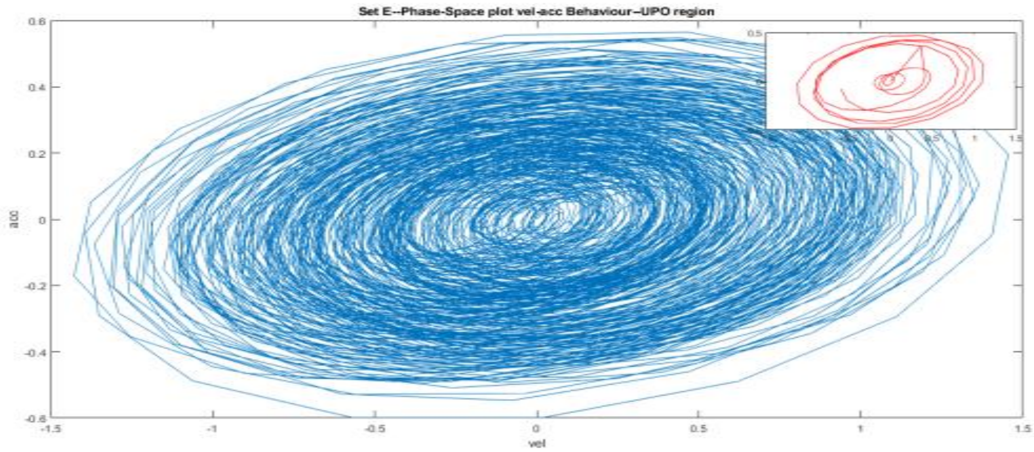


Fig 3(b)- Phase Space plot of velocity acceleration for the patient during seizure of the UPO region shows bi-stability with denser and wider trajectories following a linear harmonic motion at the centre then the extreme orbits show grazing bifurcation.

**Fig 3:** Velocity-acceleration phase space plot for UPO region for both healthy and patients during seizure.

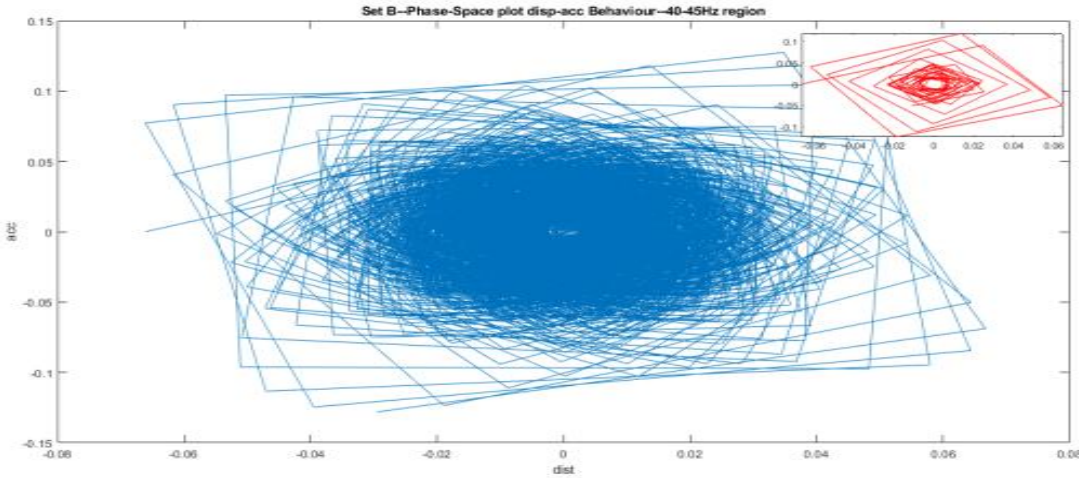


Fig 4(a): Displacement acceleration phase space plot for healthy eye closed at 40-45Hz region shows change in acceleration abruptly about four times in each loop, giving the changing loop a quadrilateral shape. The central dense region is obeying linear motion but the extreme orbits are possessing nonlinear motion like impact oscillator. This random phase change of the periodic orbit indicates the periodicity of the occurrence for healthy subject.

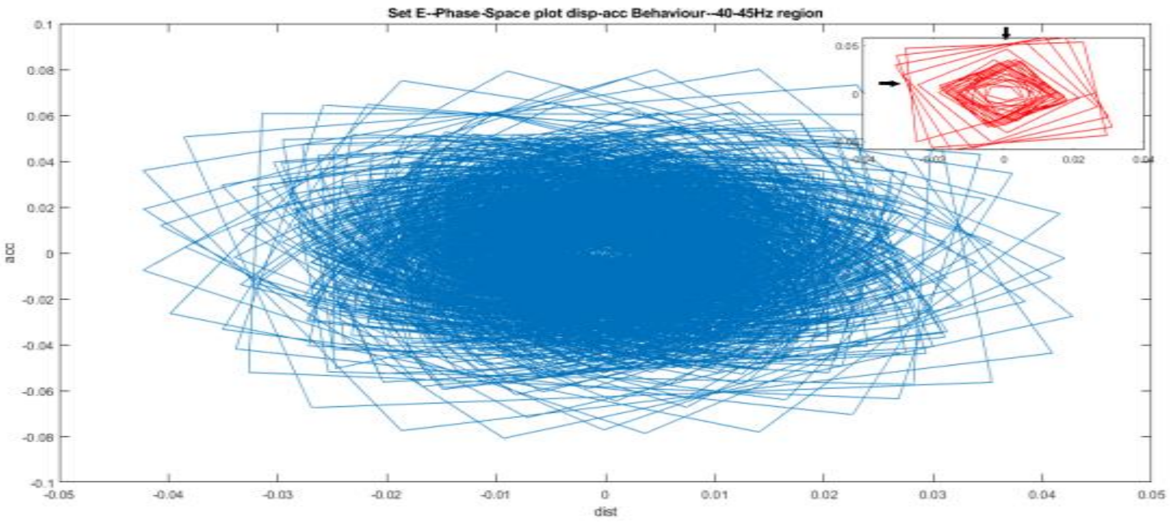


Fig 4(b): Displacement acceleration phase space behavior of the patient during seizure at 40-45Hz region separating the higher structures from the lower ones. The periodicity is from linear to nonlinear behavior at the extreme region showing piecewise linear motion giving rise to ‘lotus’ structure, from where the sudden abrupt changes of acceleration can be observed clearly due to small change of the displacement. Perhaps it indicates the state change from conscious to unconscious brain.

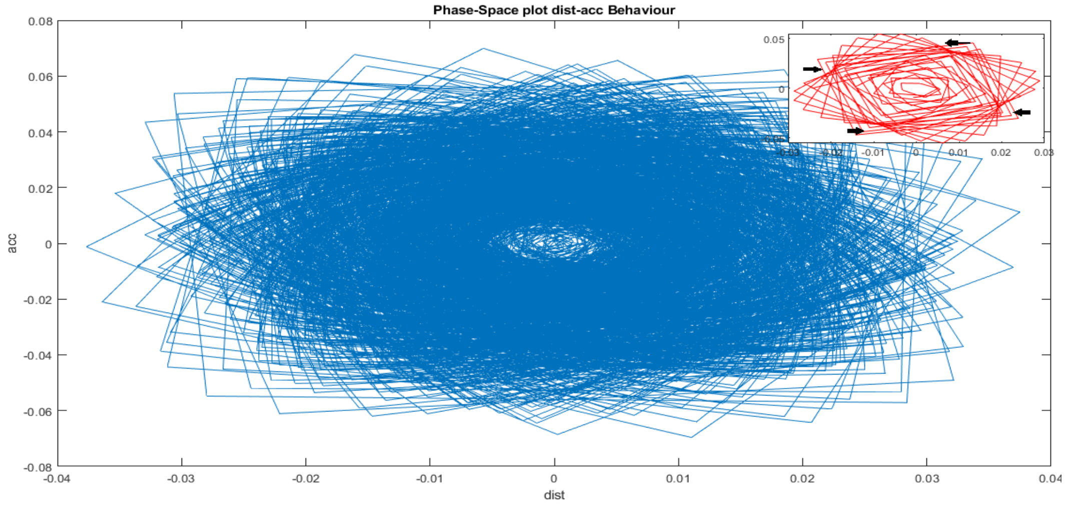


Fig 4(c): Displacement acceleration phase space structure from the epileptogenic zone of the patients (Set D) at 40-45Hz region showing a similar behavior as like the patients during seizure however here the acceleration is more evenly spread over the structure and has abrupt variation manifesting much more potential energy as compared to fig 4(b).

**Fig 4:** Displacement acceleration phase space behavior for 40-45Hz region for both healthy and patient with seizure.

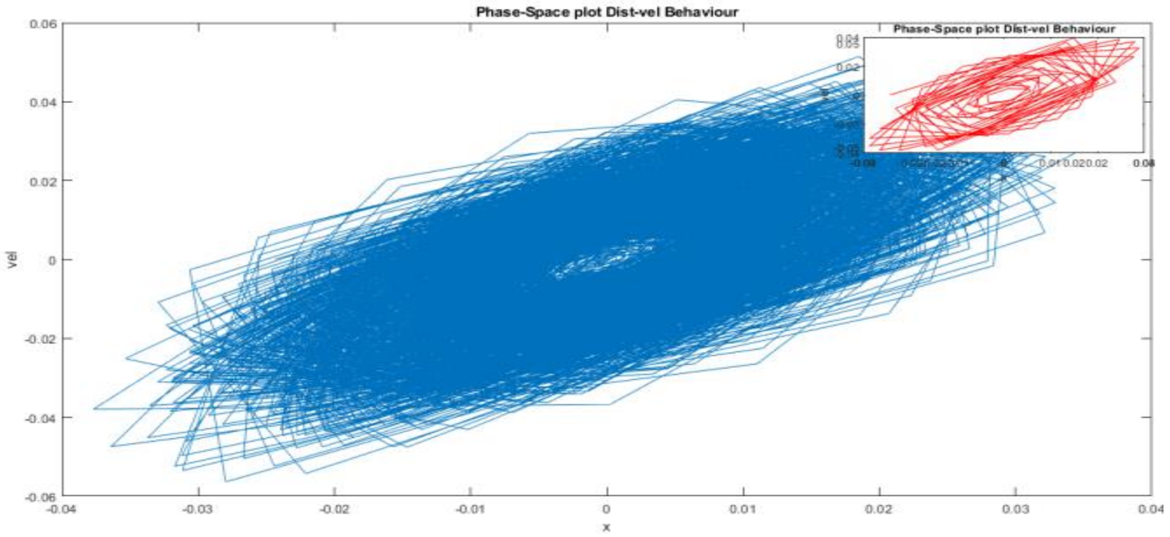


**Fig 5:** Phase space plot of displacement and velocity at 40-45Hz region for the patient from epileptogenic zone (Set D) shows the outer orbits of the concentric loops following piecewise linear motion. Here the trajectory density is more compared other figs as in 4(b) and 4(c) showing the highest contribution of the potential energy.

We have randomly chosen four other channels for the representation purpose to verify the unstable periodic orbit from the Fourier power spectrum. For Set B healthy eyes closed, we have taken the Channels O018, O043, O068 and O097 for the analysis.

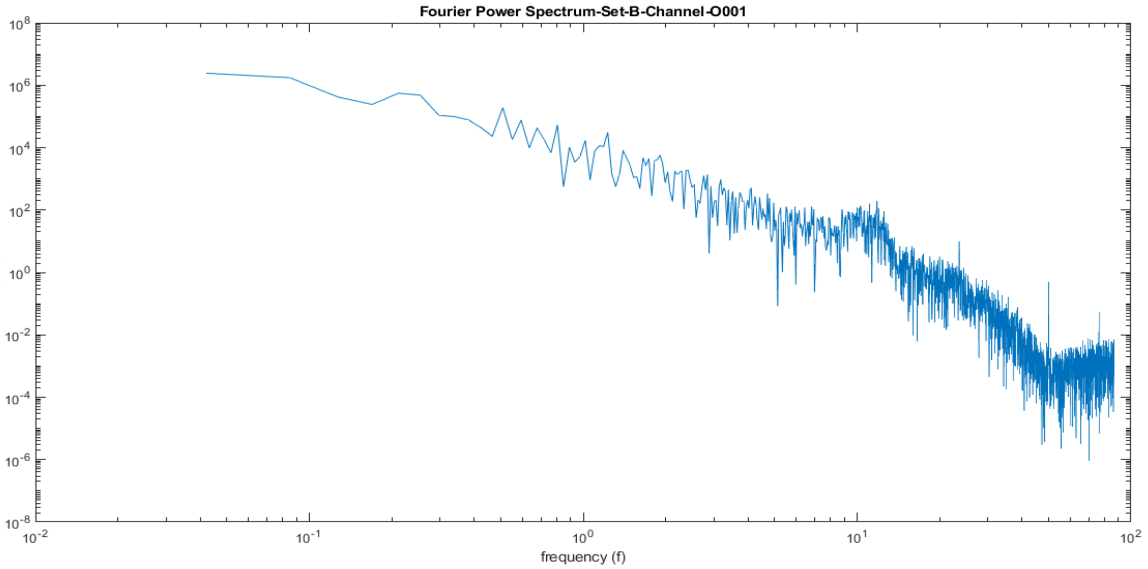

Fig:6(a): Fourier Power Spectrum of the Channel O018 of the Set B healthy eyes closed, showing the UPO at 8-14 Hz section.

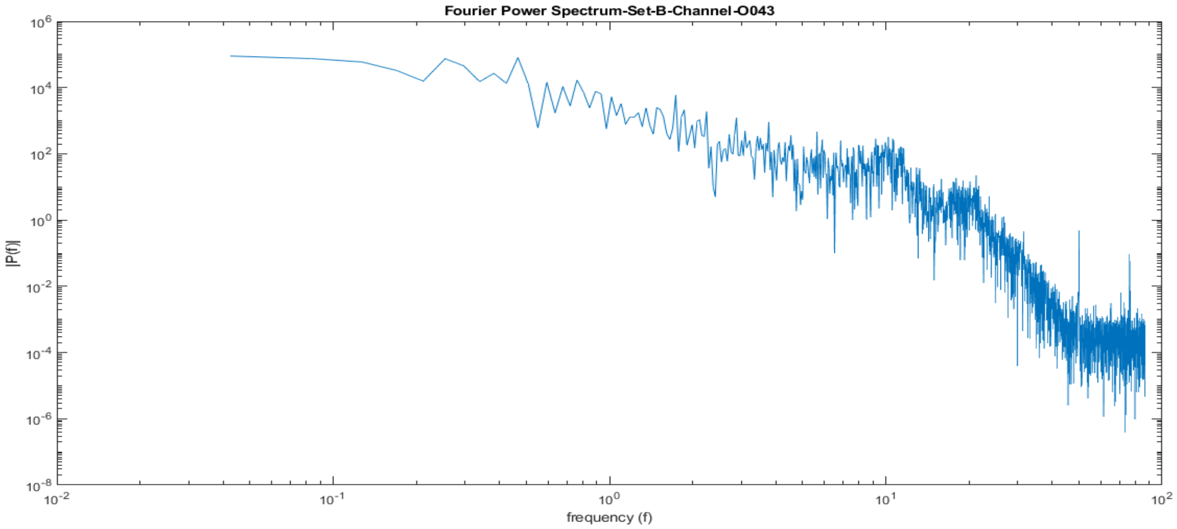


Fig:6(b): Fourier Power Spectrum of the Channel O043 of the Set B, manifesting the UPO at 8-14 Hz section, revealing its peak at around the middle (10-11Hz) of the region.

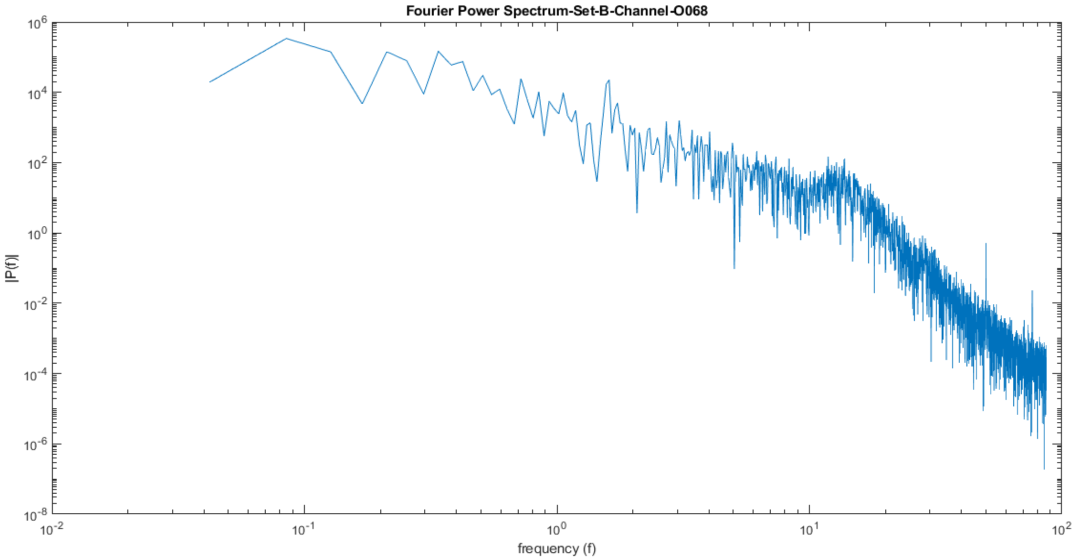


Fig:6(c): Fourier Power Spectrum of the Channel O068 of the Set B, showing the UPO range within 8-14Hz area.

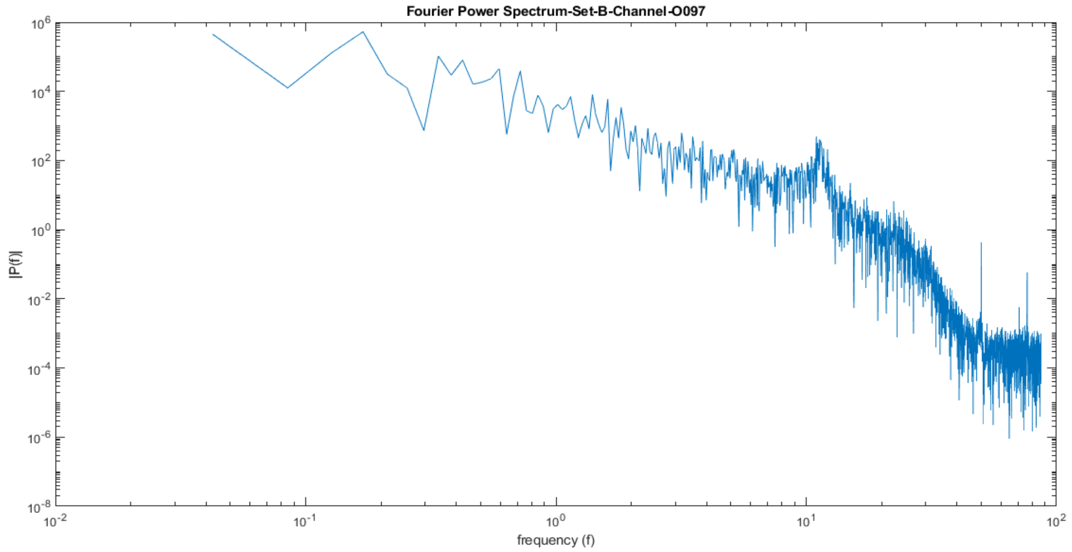


Fig:6(d): Fourier Power Spectrum of the channel O097 for Set B, with the UPO range within 8-14Hz region with the peak around 11Hz.

**Fig 6:** The Fourier Power Spectrum (FPS) of the channel O018, O043, O068, O097 of the Set B (Healthy individual with eyes close condition)

From the Fig.6, it can be observed that for healthy subjects the highest power peak is observed at the middle of the UPO region (8-14Hz) i.e. in and around 11Hz.

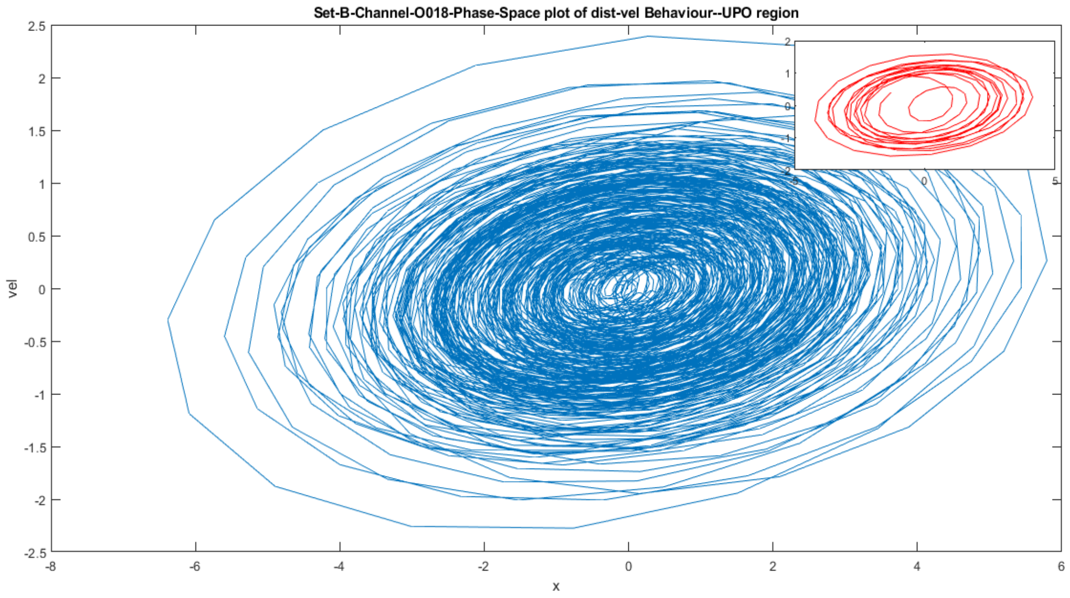


Fig 7(a): Displacement-velocity phase space for channel O018 recording of the Set B at the UPO region, showing multi periodic orbits with bi-stability as shown in the manuscript.

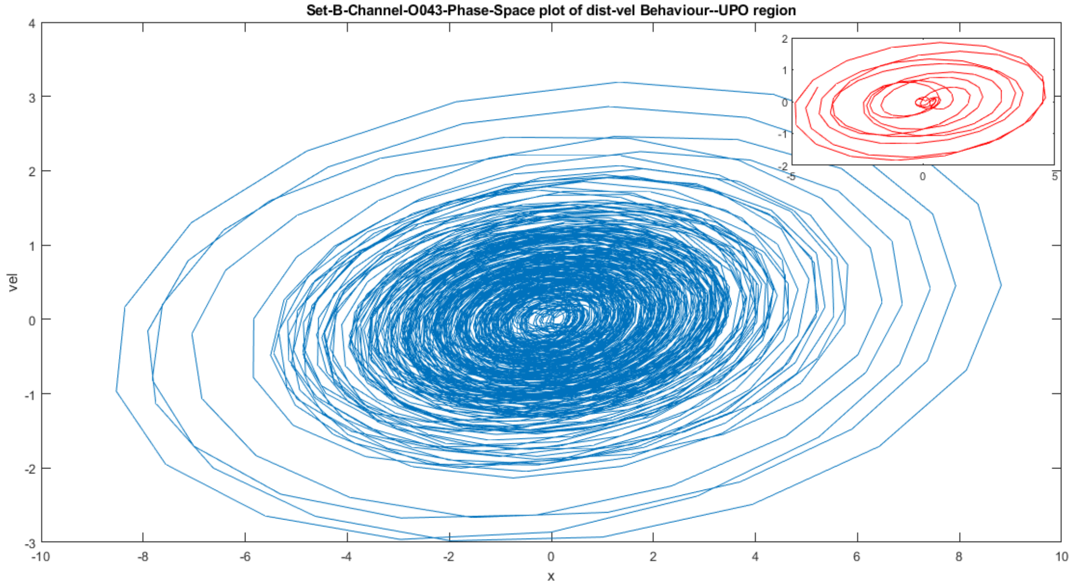


Fig 7(b): The displacement-velocity phase space behaviour of the channel O043 recording of Set B for the UPO region, showing the concentric loops where from the inset in the figure, the bi-stability behaviour is clearly observed.

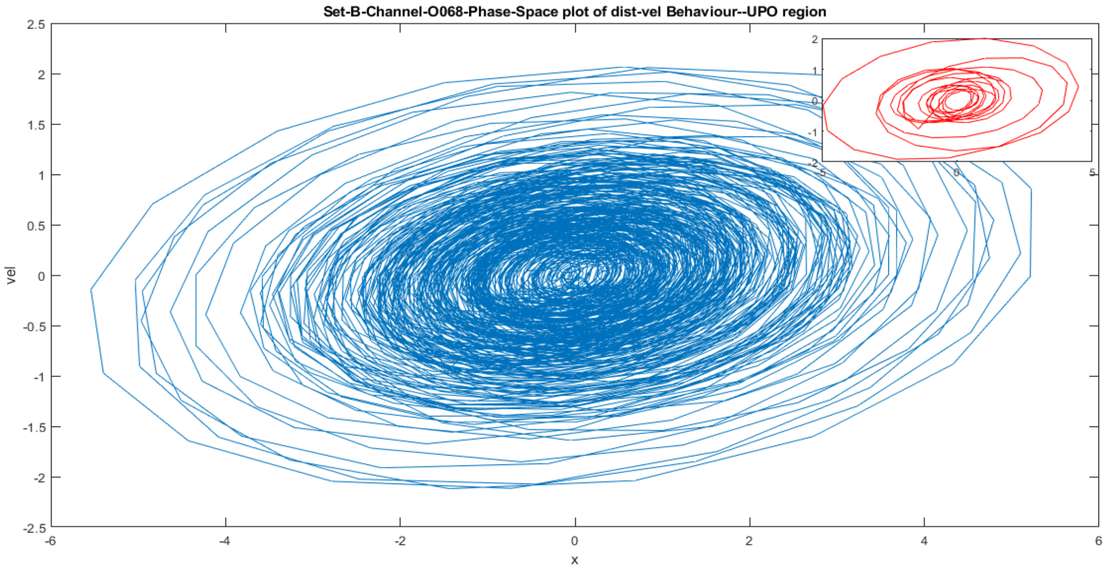


Fig 7(c): The displacement-velocity behaviour of the Channel O068 recording of the Set B for the UPO region, showing multi-periodic behaviour with number of loops of varying distance from the centre.

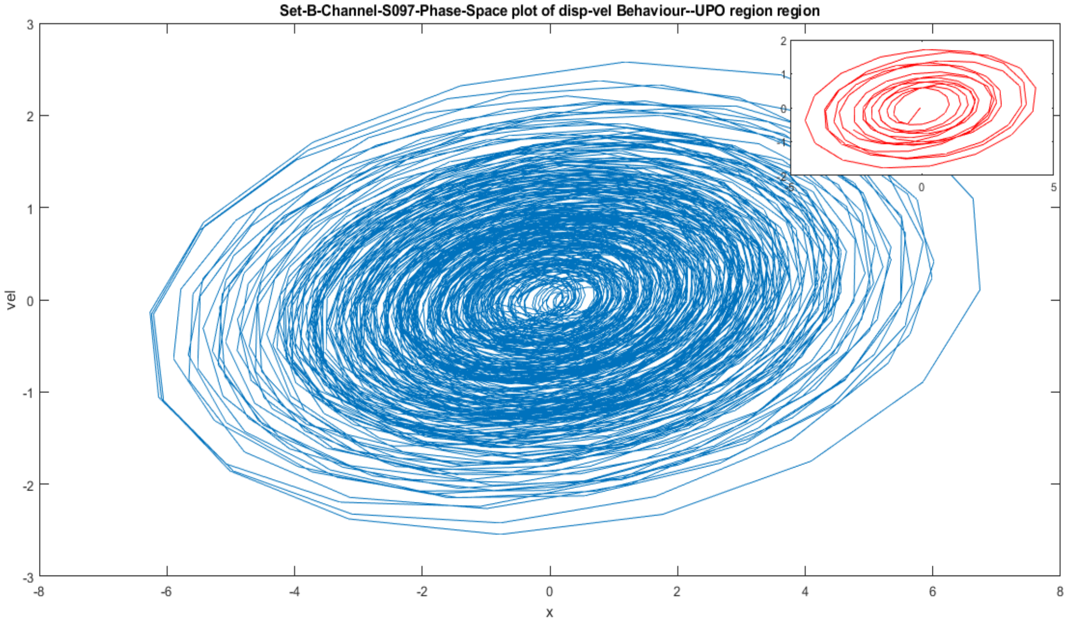


Fig 7(d): Phase-space plot of the displacement-velocity behaviour of the Channel O097 recording of the Set B for the UPO region, with a number of loops which are centrally dense and is obeying a periodic motion in the central region then slowly intending to the unbound motion.

**Fig 7:** The Phase space plot of the displacement-velocity behaviour of the Channels O018, O043, O068, O097 of the Set B for the Unstable Periodic Region (8-14Hz)

From fig 7, the phase space plots of the UPO region for the Set B, shows the bistable behaviour, as well as the linear harmonic motion in the central portion and gradually engulfing the nonlinear motion till the outer orbits, which shows grazing bifurcation leading a non impacting periodic orbit to bifurcate into the impacting one.

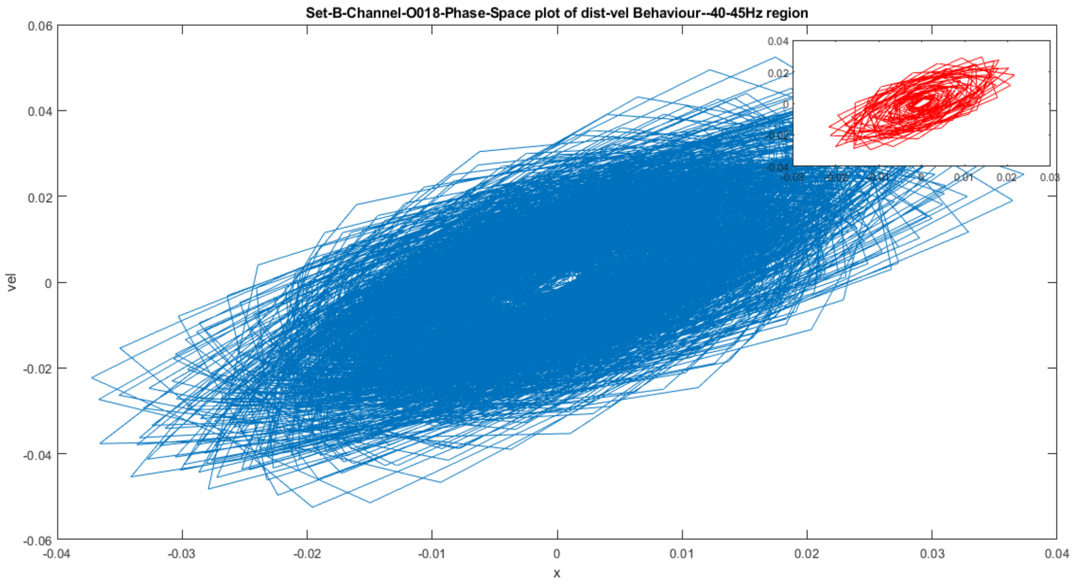


Fig 8(a): The phase-space plot of displacement and velocity for 40-45Hz region of channel O018 Set B, showing sharp changes of the acceleration with the velocities at the extreme points.

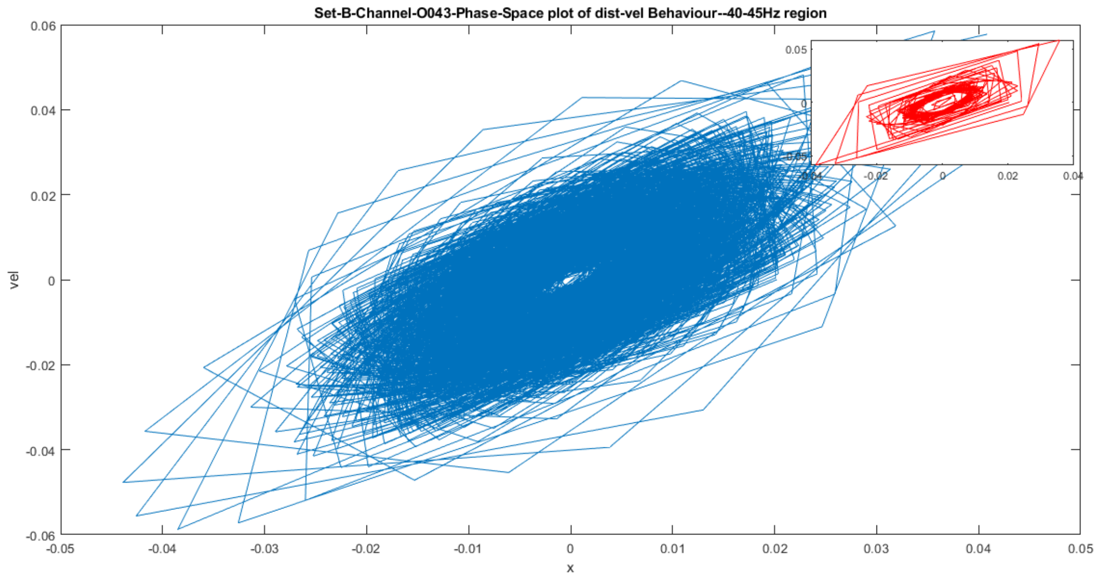


Fig 8(b): The phase-space plot of displacement-velocity for 40-45Hz region of channel O043 Set B, following a linear motion in the central region then nonlinear motion in the outer regions.

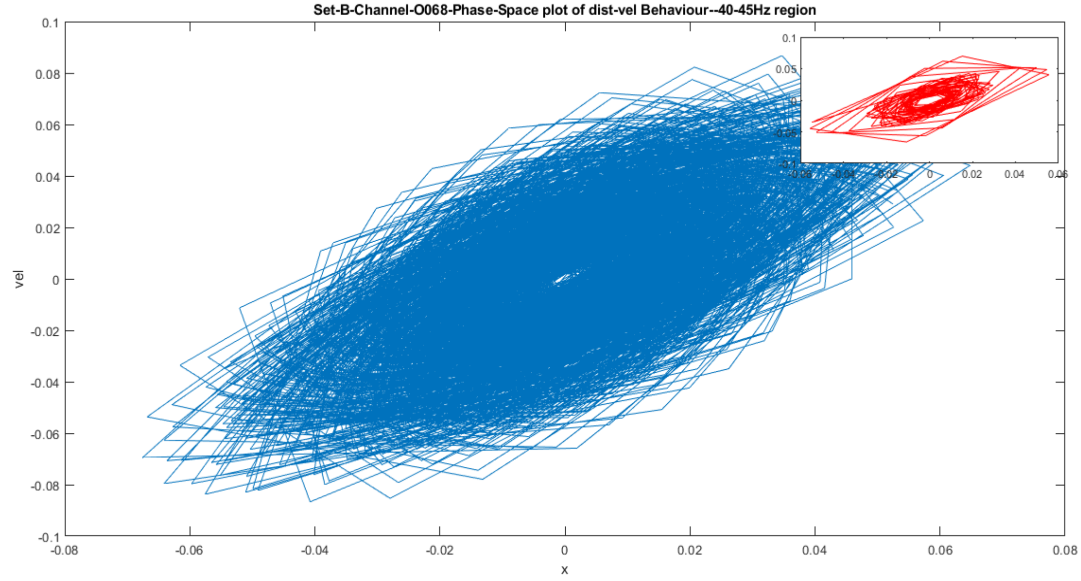


Fig 8(c): The phase-space plot of displacement-velocity for 40-45Hz region of channel O068 Set B.

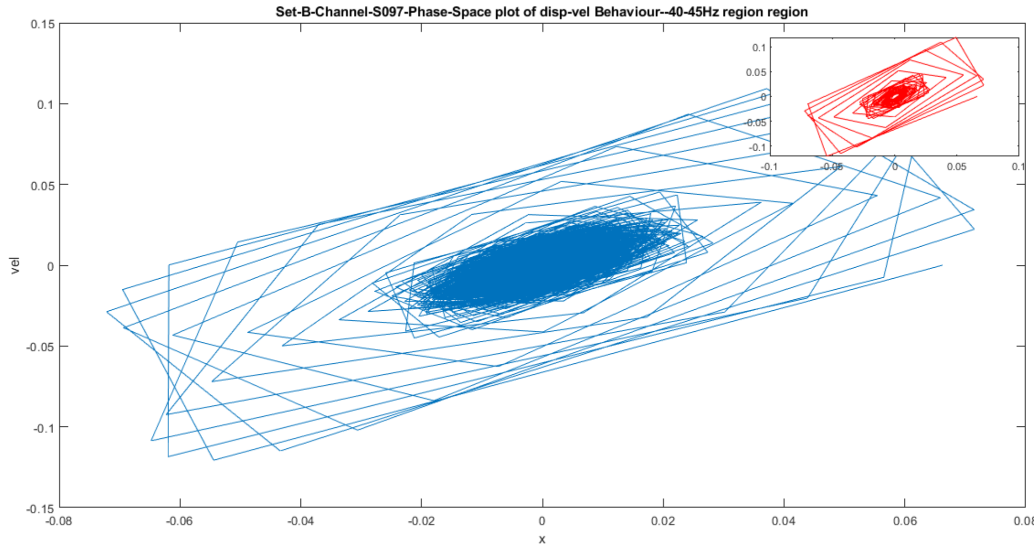


Fig 8(d): The phase-space plot of displacement-velocity for 40-45 Hz region of channel O097 Set B, showing piecewise linear motion in outer region.

**Fig 8:** The Phase space plot of the displacement-velocity behaviour of channels O018, O043, O068, O097 of Set B for the 40-45 Hz region.

Thus Fig 8 shows abrupt acceleration value changes with sharp changes at the edges. Thus a small perturbation is impacting large behavioural change over the phase space plot which we could use as a Bio-marker for the onset of the epileptic seizure.

Similarly, we have randomly chosen four other channels for the representation purpose to verify the unstable periodic orbit from the Fourier power spectrum for Set E patient during seizure. We have taken the Channels S028, S051, S076, and S092 for the analysis.

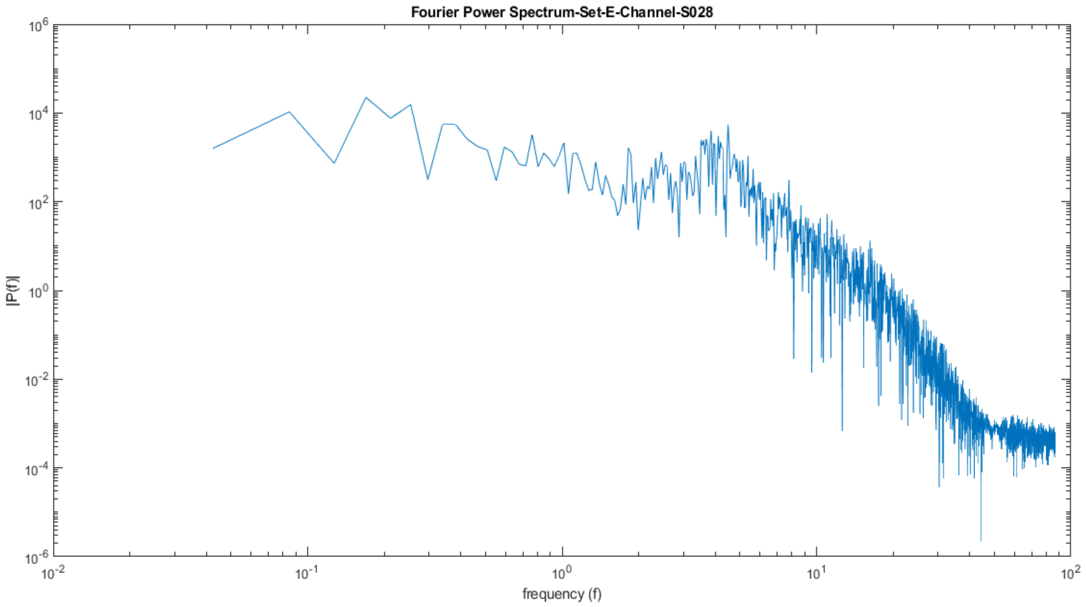


Fig 9(a)- The Fourier Power Spectrum of the Channel S028 Set E, showing the diminished UPO region in 8-14Hz section.

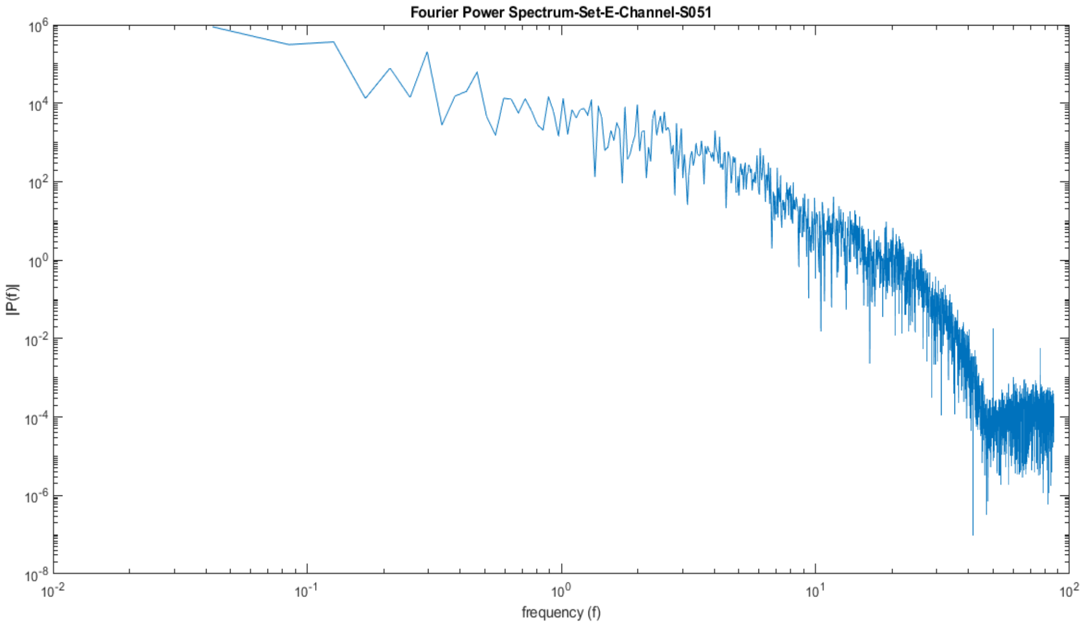


Fig 9(b) - The Fourier Power Spectrum of the Channel S051 for Set E, with no UPO peak in this UPO region

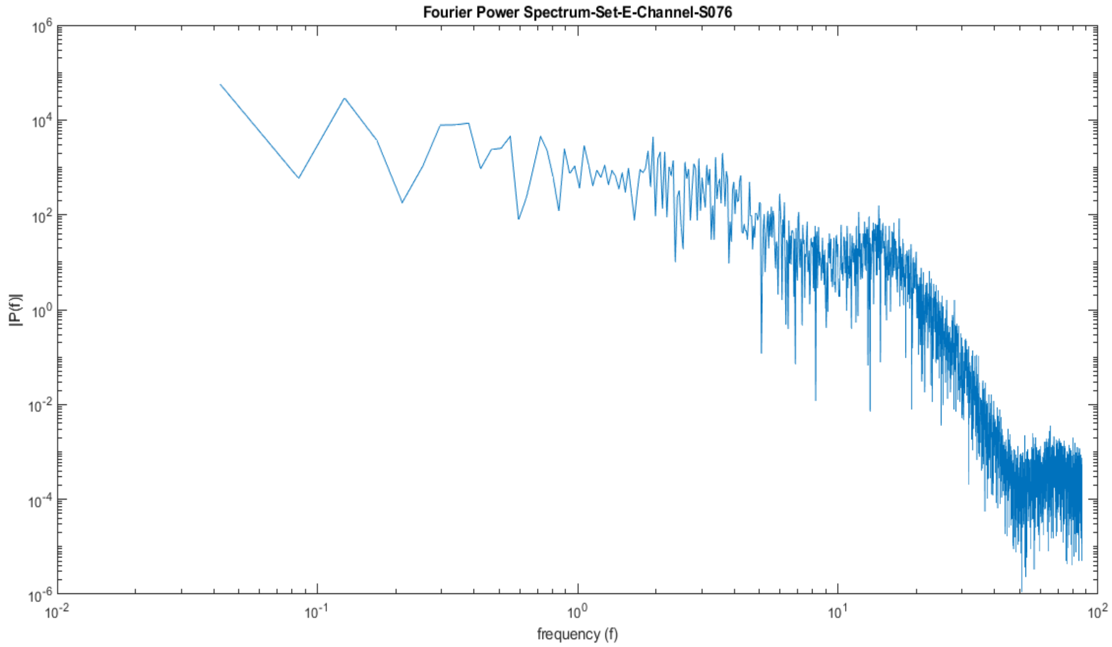


Fig 9(c) - The Fourier Power Spectrum of the Channel S076 for the Set E

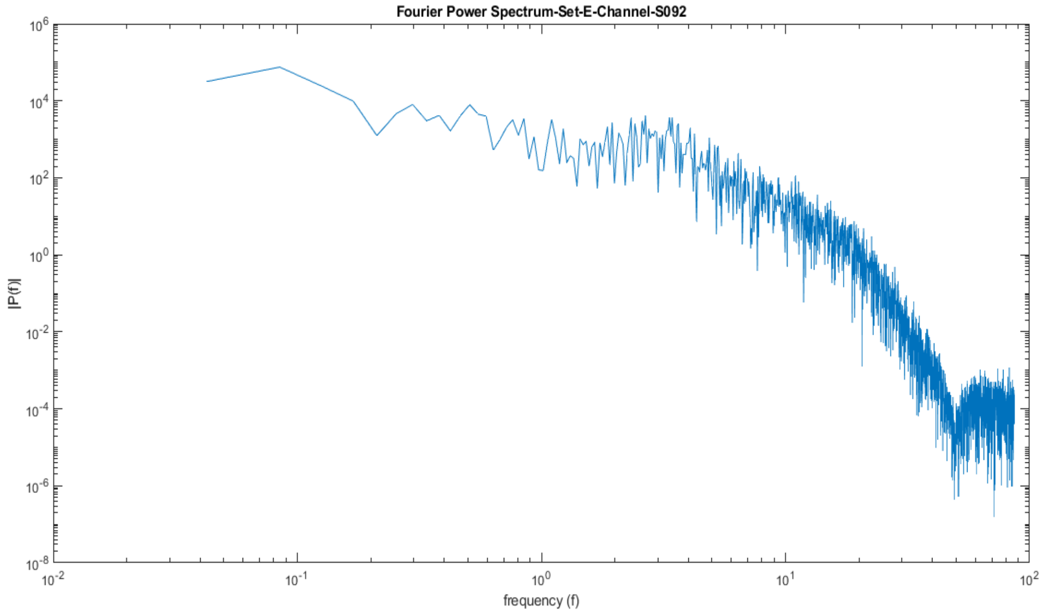


Fig 9 (d) - The Fourier Power Spectrum of the Channel-S092 for the Set E, with the diminished UPO region having peak at the first extreme point of the region.

**Fig 9:** The Fourier Power Spectrum (FPS) of the Channel S028, S051, S076, S092 of the Set E (Patient individual during epilepsy seizure)

The Fourier Power Spectrum plot for the Set E various channels showing the diminished UPO region which is the characteristic difference from the healthy subjects having the peak power emerging at either of the extreme points of the UPO region. The observed diminished power with one-sided lobe is used in our study as bio-marker for the epileptic seizure.

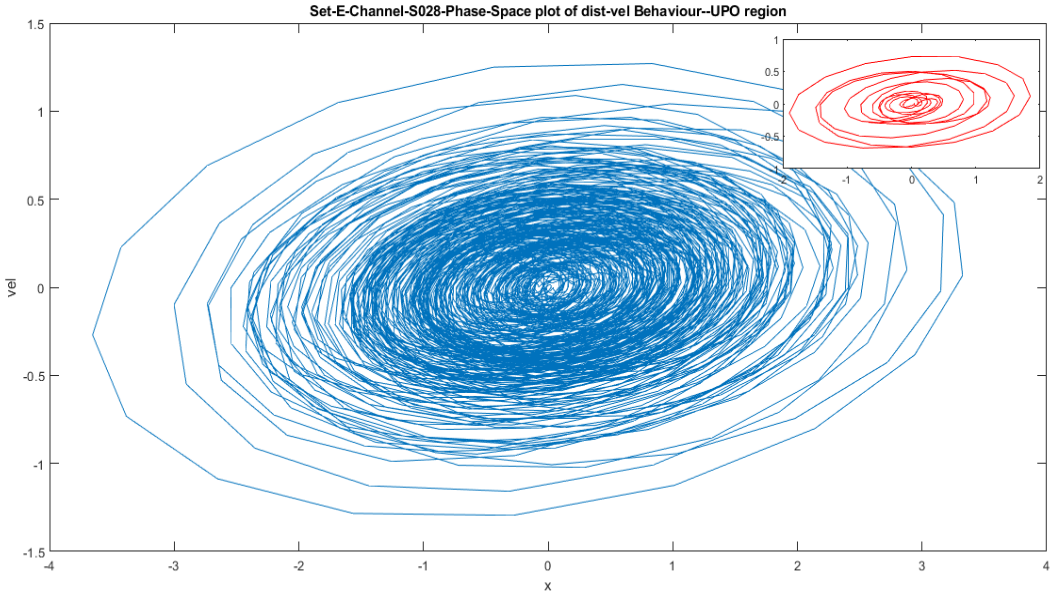


Fig 10(a)- The Phase-space plot of the channel S028 Set E for the UPO region, showing the bistable behaviour with multiple converging loops.

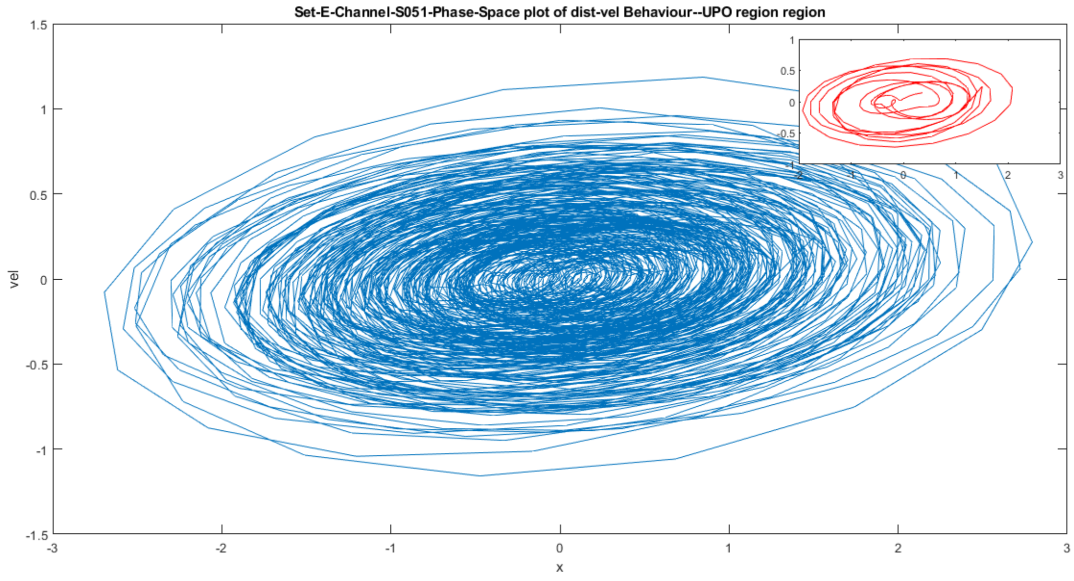


Fig 10(b)- The Phase- space plot of the channel S051 of the Set E for the UPO region, showing many converging loops with centrally linear harmonic motion.

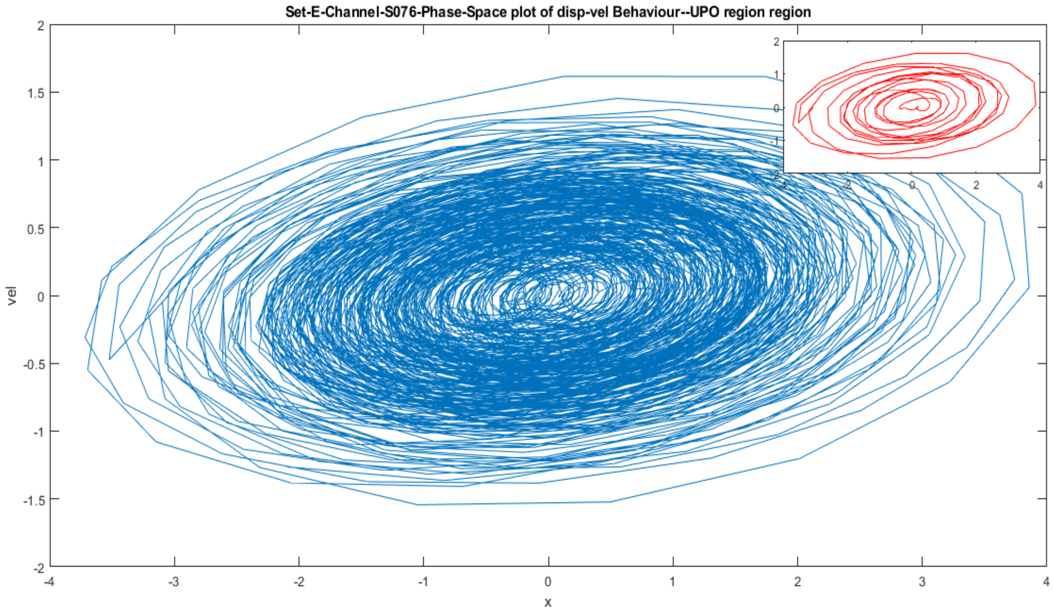


Fig 10(c)- The Phase- space plot of the channel S076 of the Set E for the UPO region

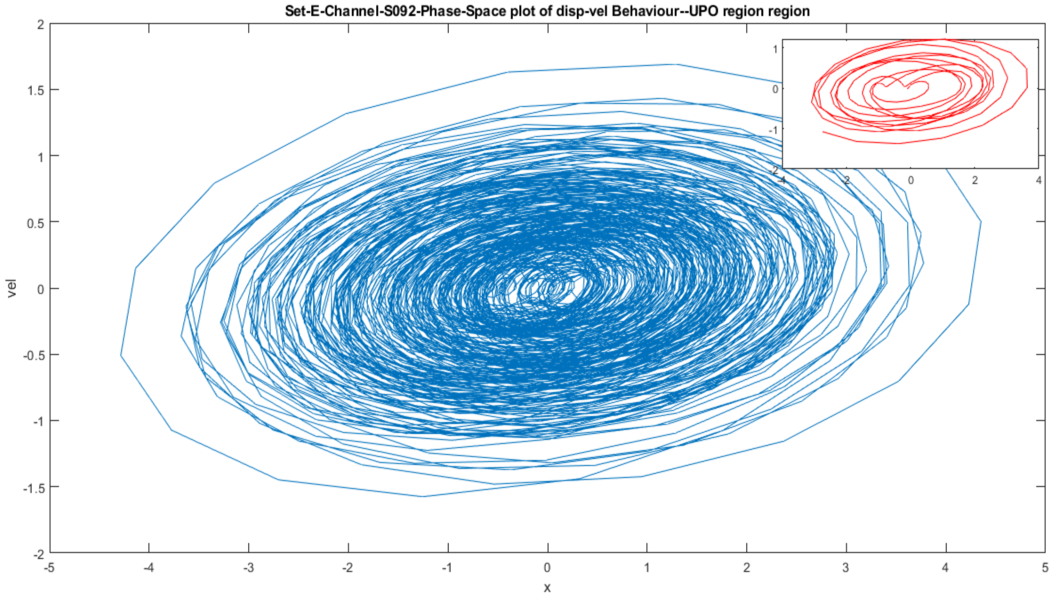


Fig 10(d)- The Phase- space plot of the channel S092 of the Set E for the UPO region, revealing oscillatory behaviour that of an impact oscillator.

**Fig 10:** The Phase space plot of the displacement-velocity behaviour of channels S028, S051, S076, S092 of Set E for the Unstable Periodic Region (8-14Hz)

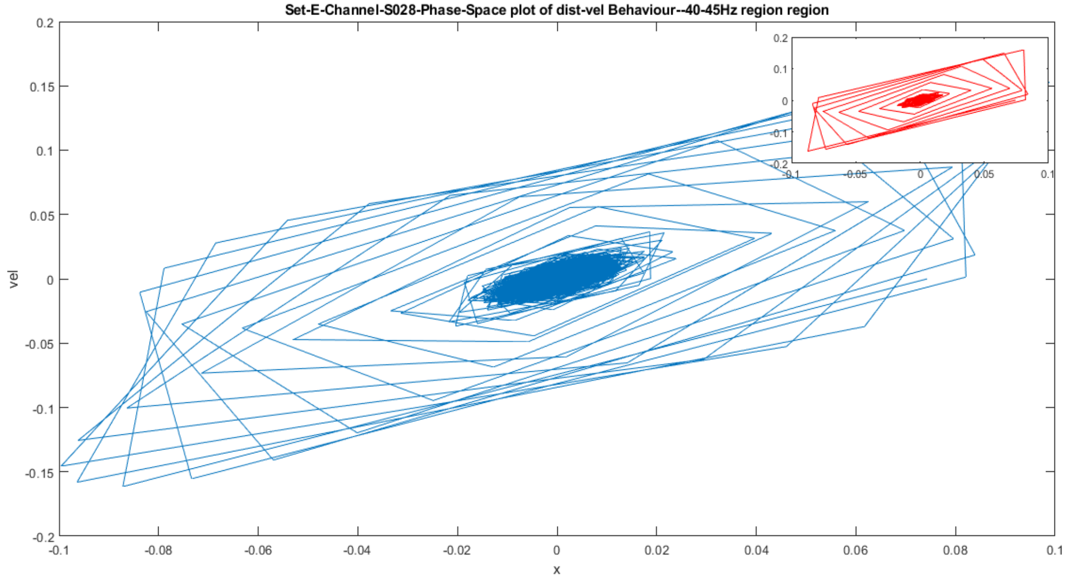


Fig 11(a): The Phase space plot of the 40-45Hz section for the Channel S028, Set E.

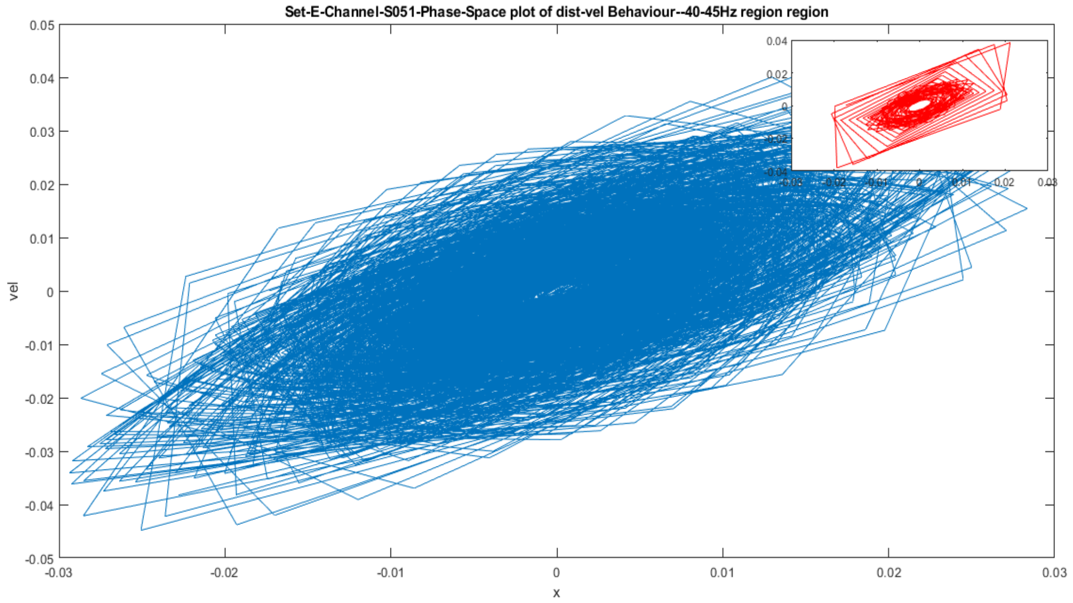


Fig 11(b): The Phase space plot of the 40-45Hz section for the Channel S051, Set E, showing a denser number of oscillatory lines and sudden abrupt changes of acceleration visible through the sharp edges of the loops.

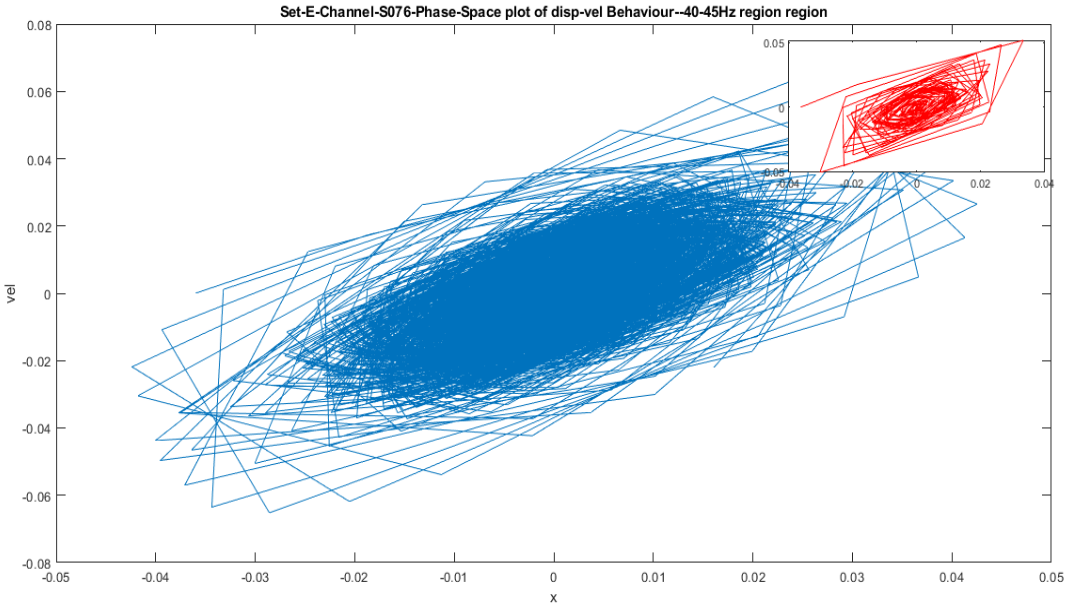


Fig 11(c): The Phase space plot of the 40-45Hz section for the Channel S076, Set E.

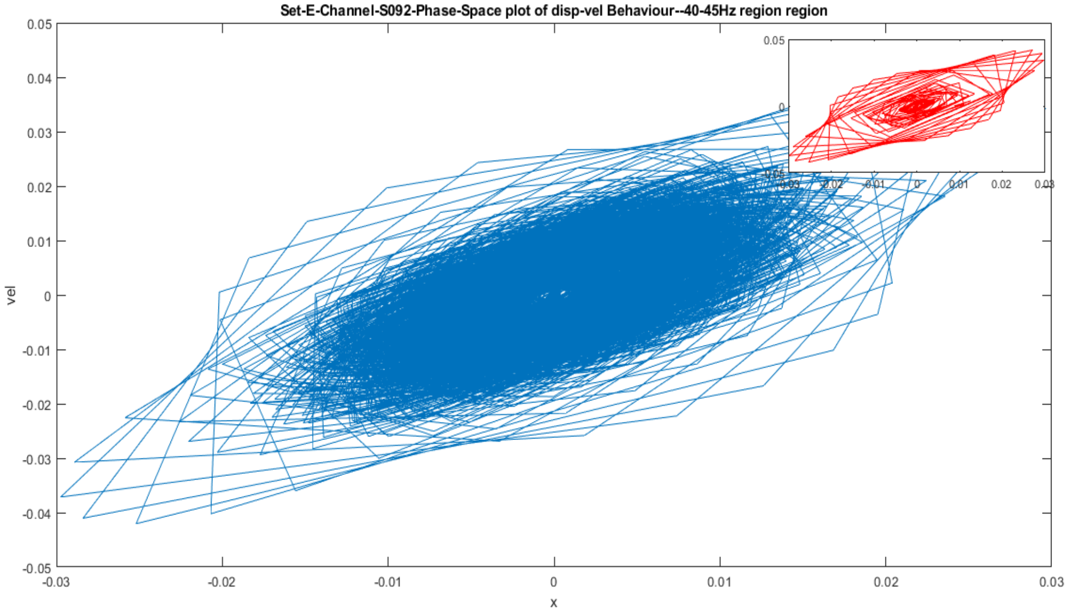


Fig 11(d): The Phase space plot of the 40-45Hz section for the Channel S092, Set E, showing piecewise linear motion in the superficial orbits following the non-linear motion.

**Fig 11:** The Phase space plot of the displacement-velocity behaviour of the Channels S028, S051, S076, S092 of the Set E for the 40-45Hz section

The Phase space plots for the UPO and 40-45Hz section for the Set E i.e. fig 10 and 11 shows denser orbits than that of healthy individual (Set B) revealing higher contribution in Set E.

### **Plots for Set A, Set C, and Set D:**

In the manuscript we discussed mainly the analysis from healthy subject during eye closed (set B) and patient during seizure (set E). Also in the supplementary material so far, we have discussed various results for Set B and E. Below we will discuss results from first channel of other datasets healthy eye open (set A), patient with no seizure from hippocampal region (set C) and patient with no seizure from epileptogenic zone (set D).

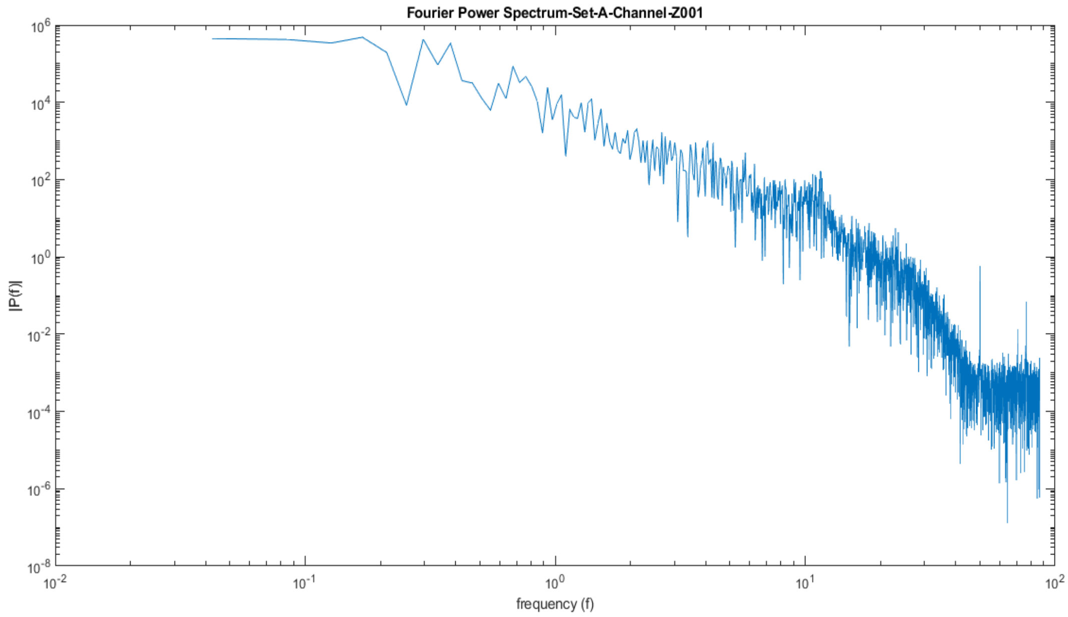


Fig 12(a)- Fourier Power Spectrum of the Set A (Channel-Z001), showing its UPO region at 8-14Hz section with its peak around 11Hz.

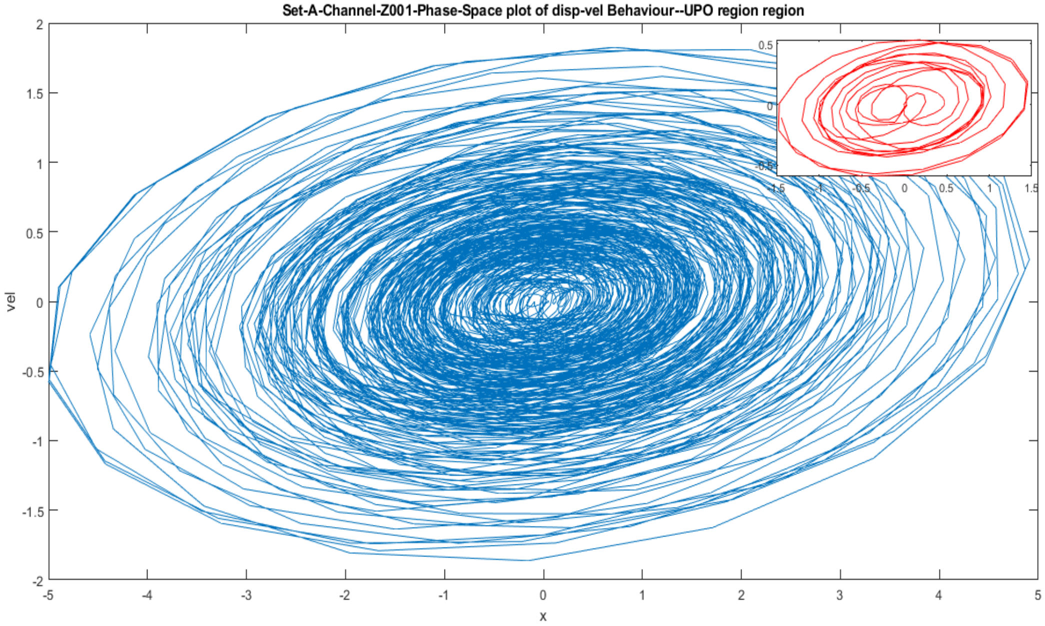


Fig 12(b)- Phase Space Plot for the displacement- velocity behaviour of the UPO region for Set A (Channel Z001) showing a single particle oscillator possessing bi-stability.

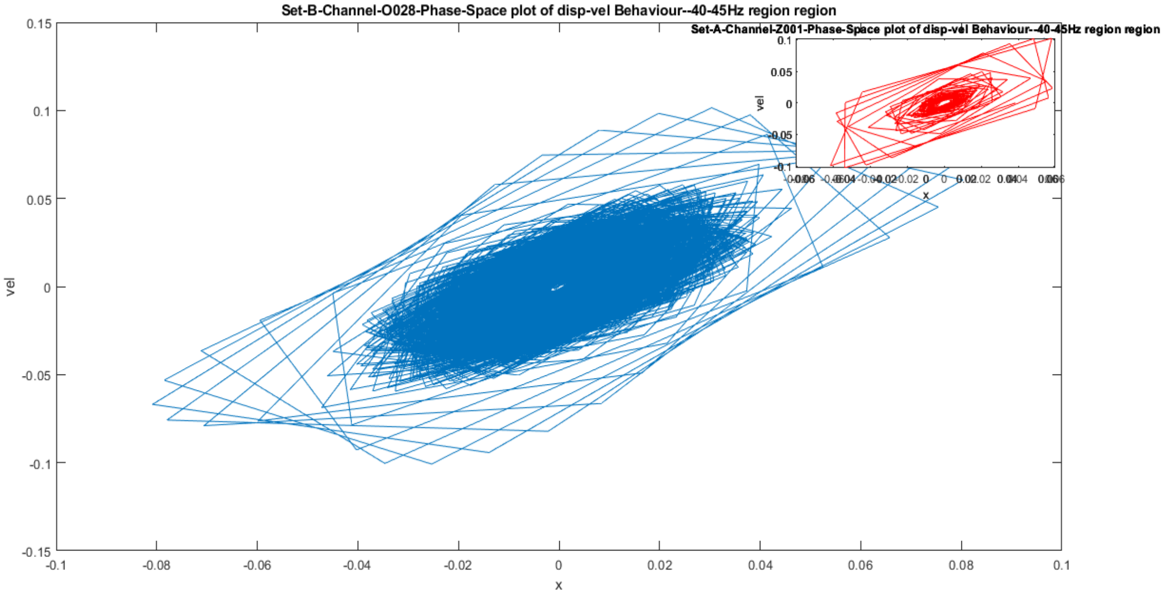


Fig 12(c)- Phase Space Plot for the displacement- velocity behaviour of the 40-45 Hz Gamma brain wave for the healthy individual with open eyes Set A (Channel Z001), manifesting linearity and nonlinearity behaviour in the central and outer region respectively.

**Fig 12-** Results of Channel-Z001, Set A EEG recording (Healthy subject with eyes open)

Set C represents the EEG data for the intracranial recording from hippocampal formation of the opposite hemisphere of the brain of patients.

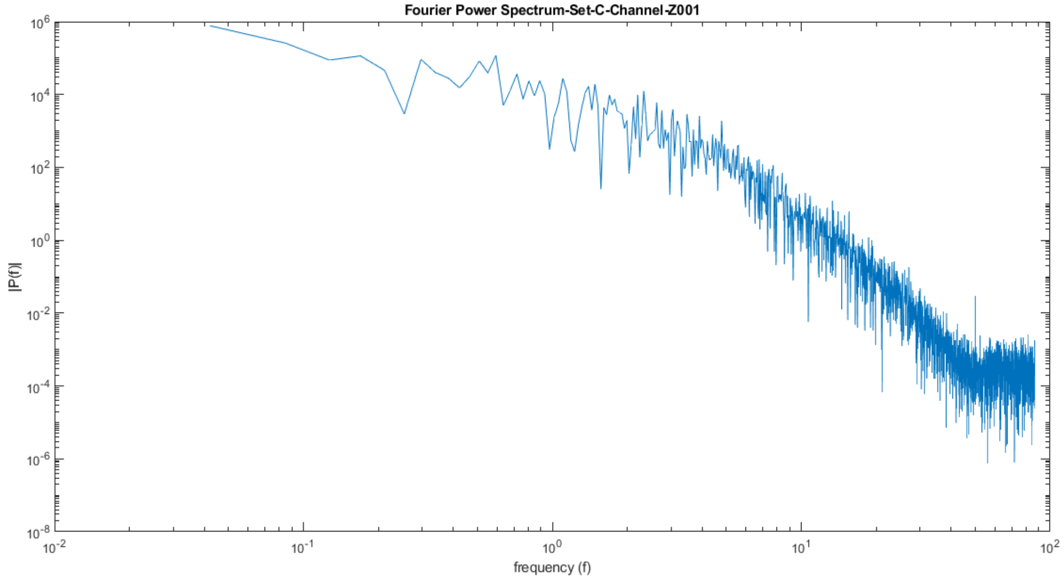


Fig 13(a)- Fourier Power Spectrum of the Set C channel (N001) showing diminished or almost non-identifiable UPO region.

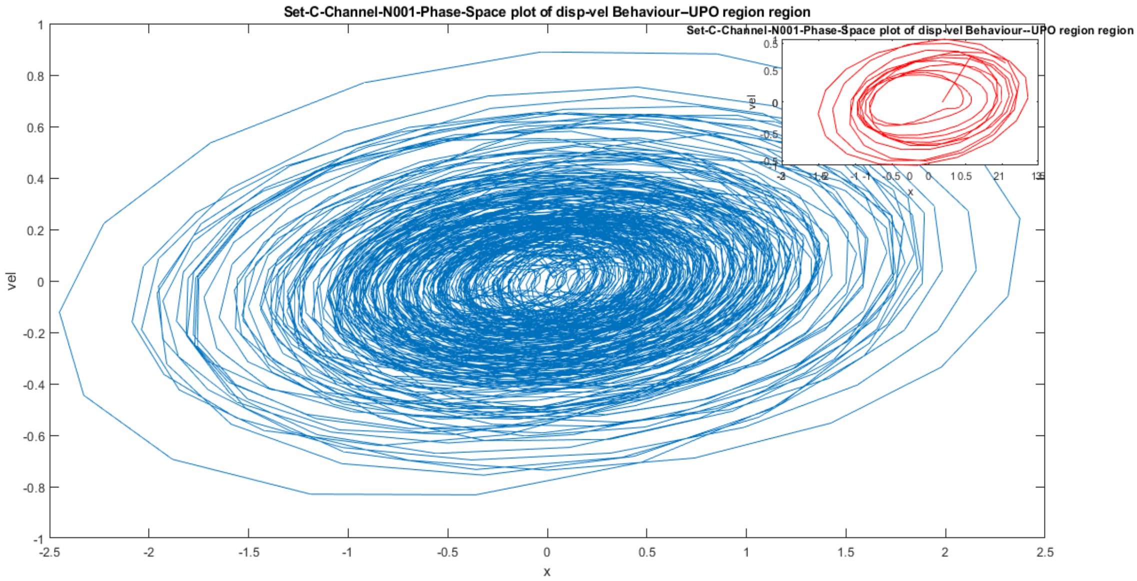


Fig 13(b)- Phase space plot of the EEG recording of the patient from Set C, channel N001, from the UPO region showing periodic motion and the extreme orbits possessing a grazing bifurcation.

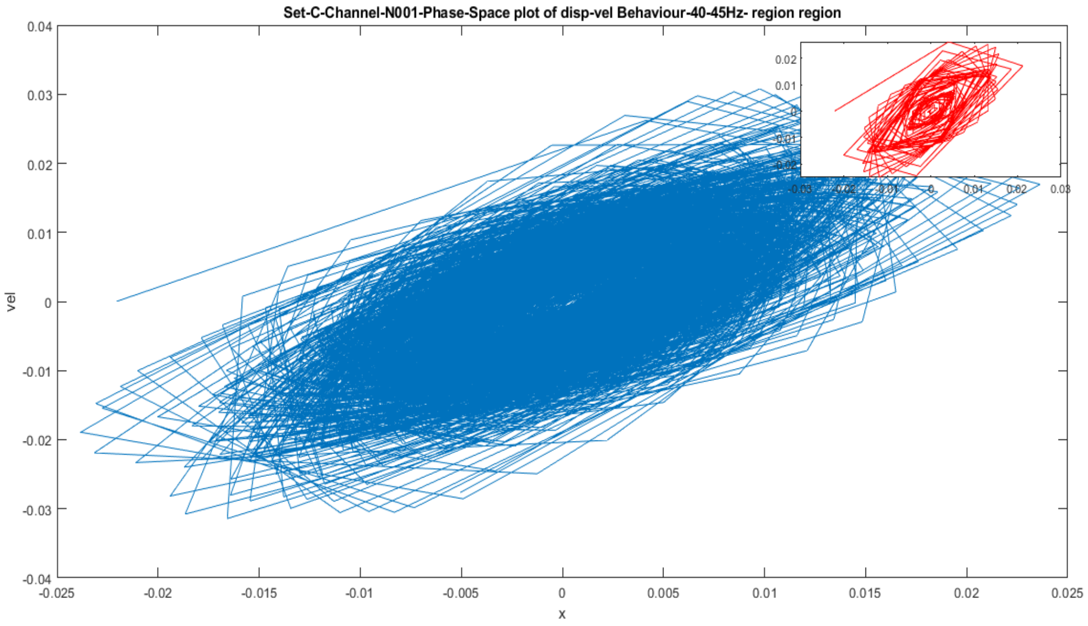


Fig 13(c)- Phase space plot of the EEG recording of the patient from Set C, channel N001, from the Gamma region 40-45 Hz, revealing its higher contribution of the potential energy through the denser orbits.

**Fig 13:** Results of the Set C (Channel N001), EEG data for the intracranial recording from patient's hippocampal formation of the opposite hemisphere of the brain.

Set D- Represents the EEG recording from within the epileptogenic zone during no seizure.

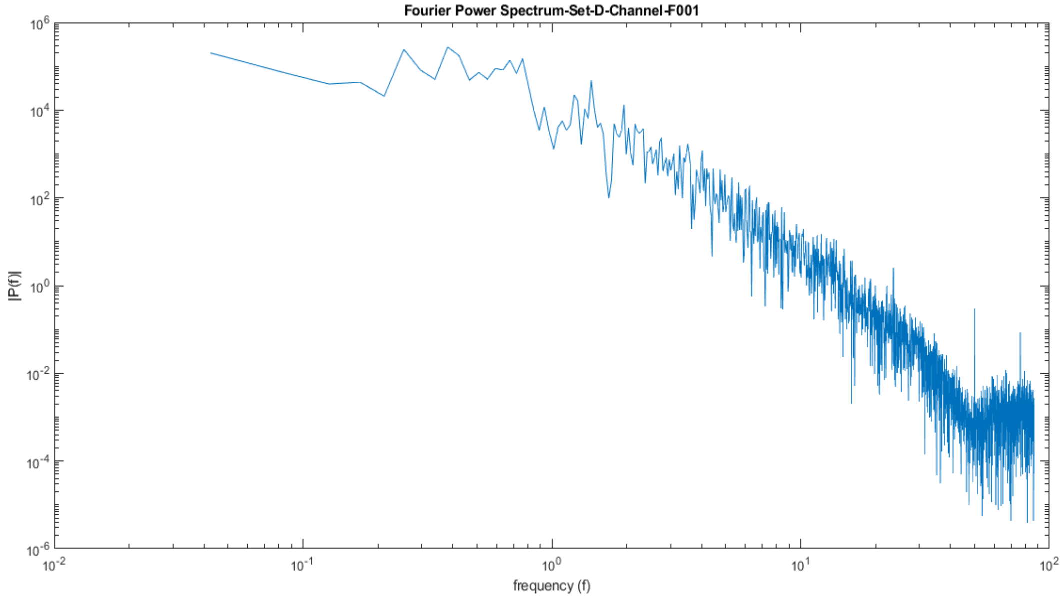


Fig 14(a)- The Fourier Power Spectrum for the Set D, within the epileptogenic zone (Channel F001), where we observe absence of UPO region.

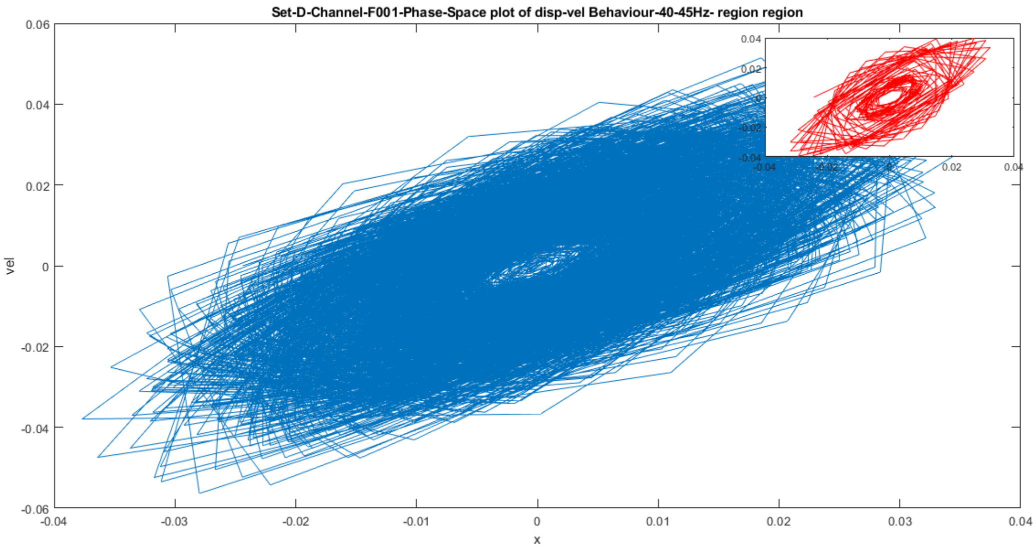


Fig 14(b)- Phase space plot for the Set D, Channel F001, showing the displacement-velocity behaviour of the gamma region 40-45Hz depicting piecewise linear motion with sharp cones. The central linear region engulfing the behaviour of nonlinearity.

**Fig 14-** Results of channel F001, Set D EEG recording within the epileptogenic zone of the epileptic patient.

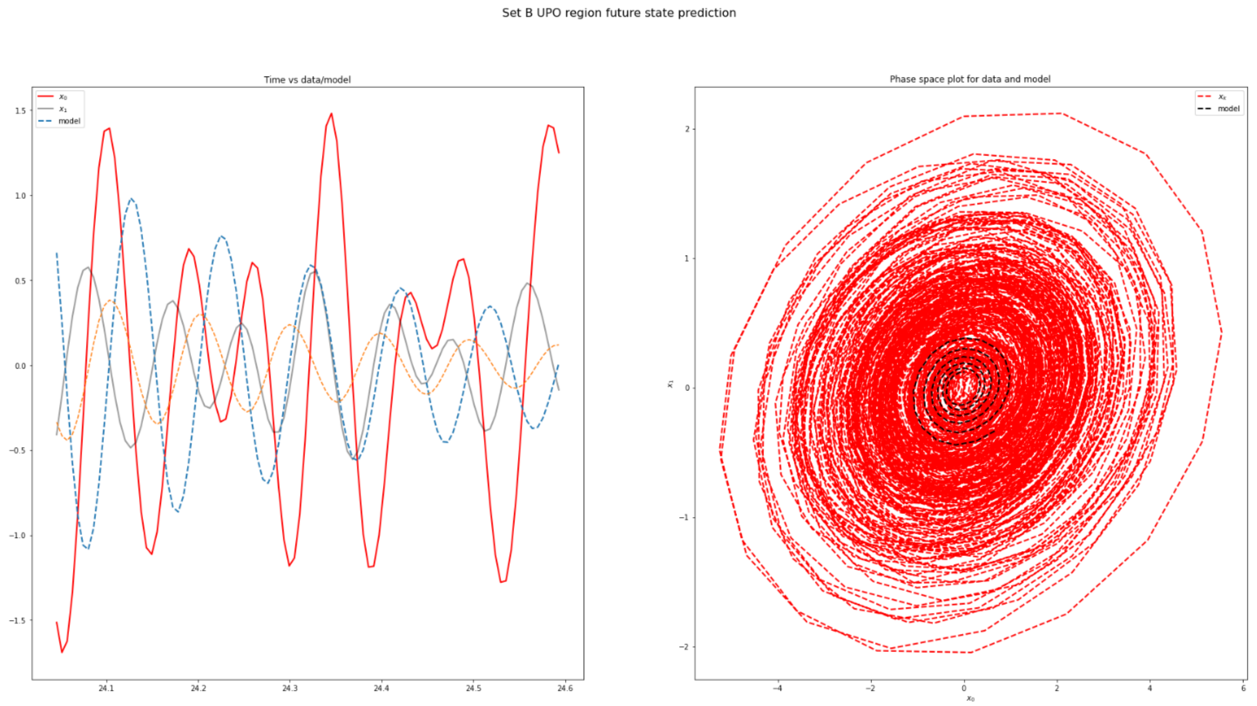


Fig 15(a)- Plot of displacement-velocity phase space data along with the model data for set B healthy eye closed UPO region. We observe the derived governing equation/model matches to the phase space displacement-velocity data.

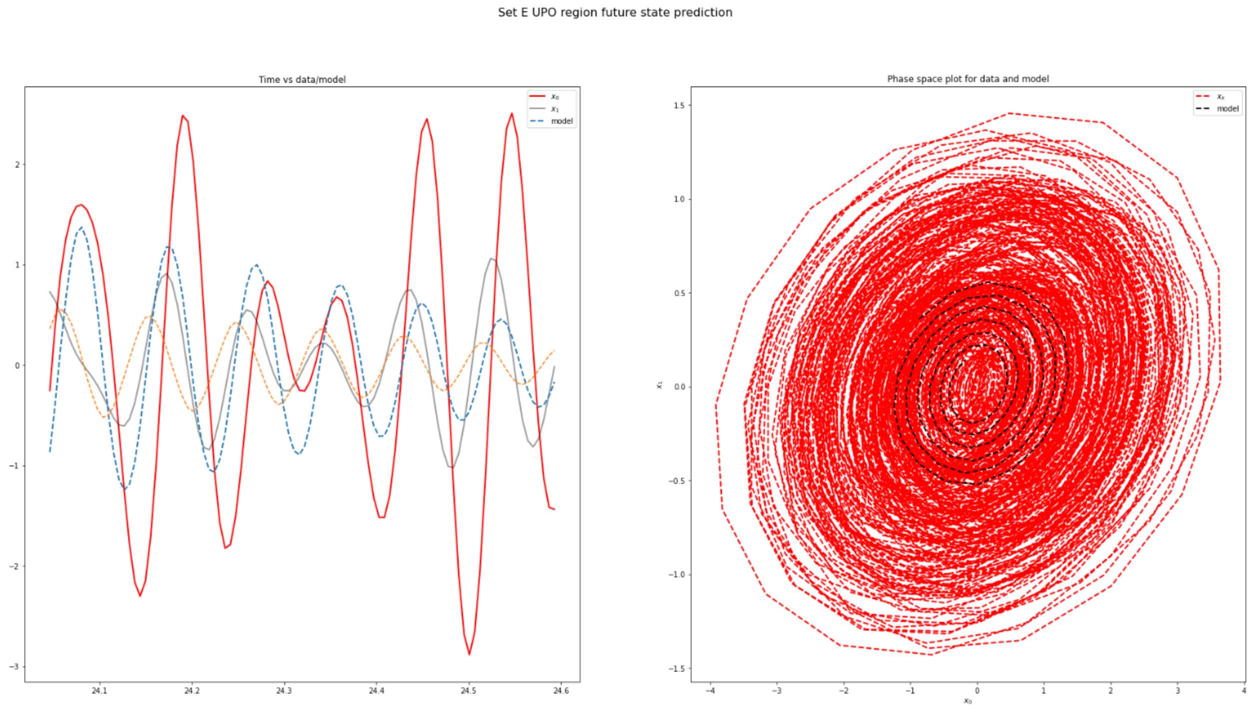


Fig 15(b)- Plot of displacement-velocity phase space data along with the model data for set E patient during seizure UPO region. We observe the derived governing equation/model matches to the phase space displacement-velocity data.

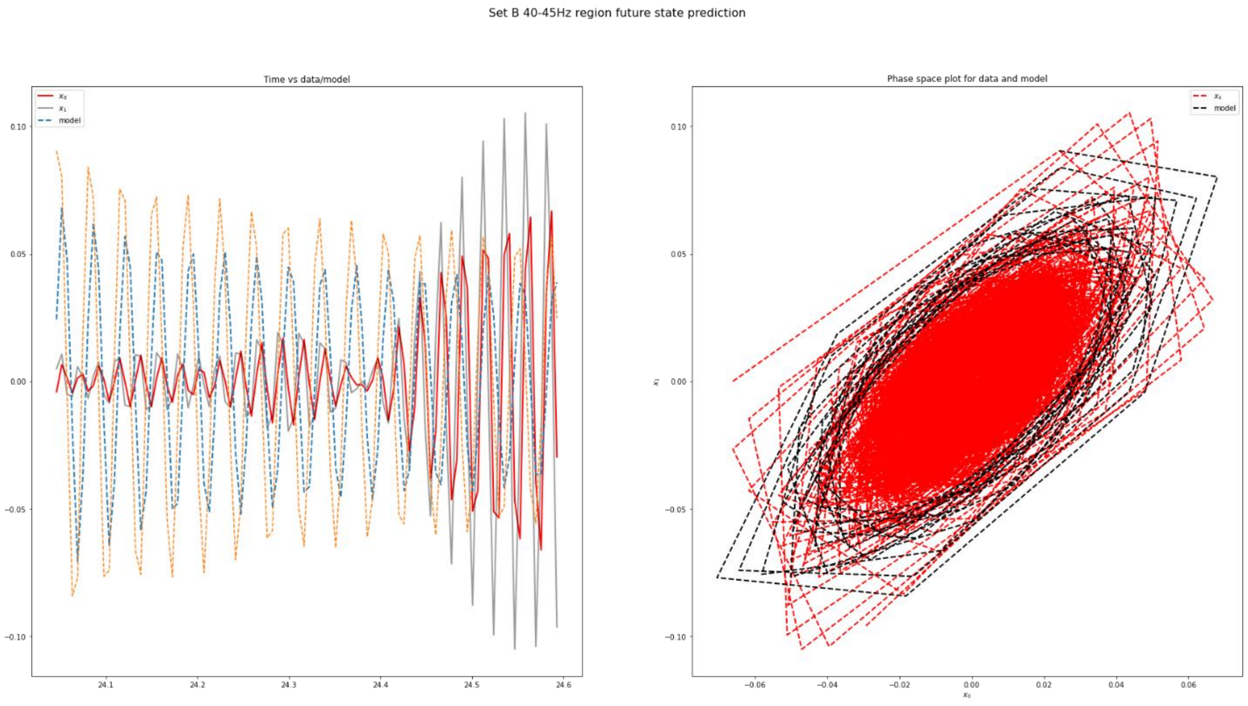


Fig 15(c)- Plot of displacement-velocity phase space data along with the model data for set B healthy during eye closed 40-45Hz region. We observe the derived governing equation/model matches to the phase space displacement-velocity data.

**Fig 15-** Plot of data and model that matches closely.
